# Disruption of the PAR3/INSC/LGN complex causes microtubule instability and peripheral neuropathy

**DOI:** 10.1101/2023.08.30.23294746

**Authors:** Jui-Yu Yeh, Hua-Chuan Chao, Yu-Chien Hung, Fei-Yang Tzou, Cheng-Tsung Hsiao, Cheng-Ta Chou, Yu-Shuen Tsai, Yi-Chu Liao, Shu-Yi Huang, Marina Kennerson, Yi-Chung Lee, Chih-Chiang Chan

## Abstract

PAR3/INSC/LGN form an evolutionarily conserved complex required for asymmetric cell division in the developing brain, but its post-developmental function and disease relevance in the peripheral nervous system (PNS) remains unknown. We mapped a new locus for axonal Charcot-Marie-Tooth disease (CMT2) and identified a missense mutation c.209T>G (p.Met70Arg) in the INSC (*INSC*) gene. Modelling the *INSC^M70R^*variant in *Drosophila*, we showed that it caused proprioceptive defects in adult flies, leading to gait defects resembling those in CMT2 patients. Cellularly, PAR3/INSC/LGN dysfunction caused tubulin aggregation and necrotic neurodegeneration, with microtubule-stabilizing agents rescuing both morphological and functional defects of the *INSC^M70R^*mutation in the PNS. Our findings underscore the critical role of the PAR3/INSC/LGN machinery in the adult PNS and highlights a potential therapeutic target for INSC-associated CMT2.

**One-Sentence Summary:** PAR3/INSC/LGN dysfunction causes peripheral neuropathy and is potentially treatable by stabilizing the microtubule network.

## Introduction

Charcot-Marie-Tooth disease (CMT) is a clinically and genetically heterogeneous group of inherited neuropathies characterized by slowly progressive distal limb weakness and atrophy, sensory loss, foot deformities, and depressed tendon reflexes (*1*). The population prevalence of CMT is estimated at 1 in 2,500 individuals (*1*). Clinically, CMT is classified into demyelinating (CMT1) or axonal (CMT2) types based on forearm motor nerve conduction velocities (MNCVs) below or above 38 m/s, respectively (*2*). Although mutations in more than 100 genes have been implicated in CMT, the genetic diagnosis for more than 70% of CMT2 patients remains elusive (*3*). Notably, several CMT2 subtypes, including CMT2D (*GARS1*), CMT2E (*NEFL*), and CMT2F (*HSPB1*), share the common defect of decreased microtubule (MT) stabilization caused by disrupting α-tubulin acetylation (*4–6*), leading to MT break down into tubulin subunits and causing axonal defects. MTs regulate axonal transport by forming a dynamic network that enables efficient intraneuronal transport. Understanding the underlying genetic and molecular mechanisms can aid in improving the diagnosis, treatment, and management of CMT and provide crucial insights into the pathogenic mechanisms of neurodegenerative disorder.

During the development of the central nervous system, asymmetric cell division plays a critical role in balancing stem cell self-renewal and differentiation in the brain (*7–9*). The PAR3(PARD3)/INSC/LGN(GPSM2)(hereafter referred as PIL) complex is crucial for asymmetric cell division (ACD), regulating MTs to establish cell polarity. Mutations of the PIL complex have been shown to cause neurodevelopmental disorders, including neural tube defects resulting from the pathogenic variants of PAR3(*10*) and Chudley-McCullough syndrome (CMS) caused by LGN variants (*11*). While LGN mutations in CMS patient cells impaired MT stability, Par3-deficient mice with neural defects exhibited unstable MT in neural progenitor cells (*12*). Despite the well-established role of PAR3 and LGN during CNS development, there is no published evidence for the requirement of the PIL complex in the adult peripheral nervous system (PNS).

Unlike PAR3 and LGN, INSC has never been linked to any genetic disorder. INSC was first identified in *Drosophila* larval neuroblasts (*7*). *Drosophila* LGN is required for INSC to asymmetrically localize in ACD (*13*). INSC and LGN participate in the cytoskeleton-membrane association in the apical side of neuroblasts and induce pulling forces on the astral microtubule for the asymmetric division (*14*). Subsequently, the dynein-adaptor protein NuMA competes with INSC for LGN binding (*15*). The LGN-bound NuMA complex can recruit dynein, a MT motor protein, to induce pulling forces on the astral microtubule for asymmetric divisions (Fig.1A) (*16*). LGN encodes an evolutionarily conserved tetratricopeptide repeat (TPR) motif that interacts with the LGN-binding domain (LBD) of INSC (Fig.1B) (*13*, *17*). Whether the PIL complex, especially INSC, may be involved in CMT pathology via its role in MT regulation is not known.

**Fig. 1.**
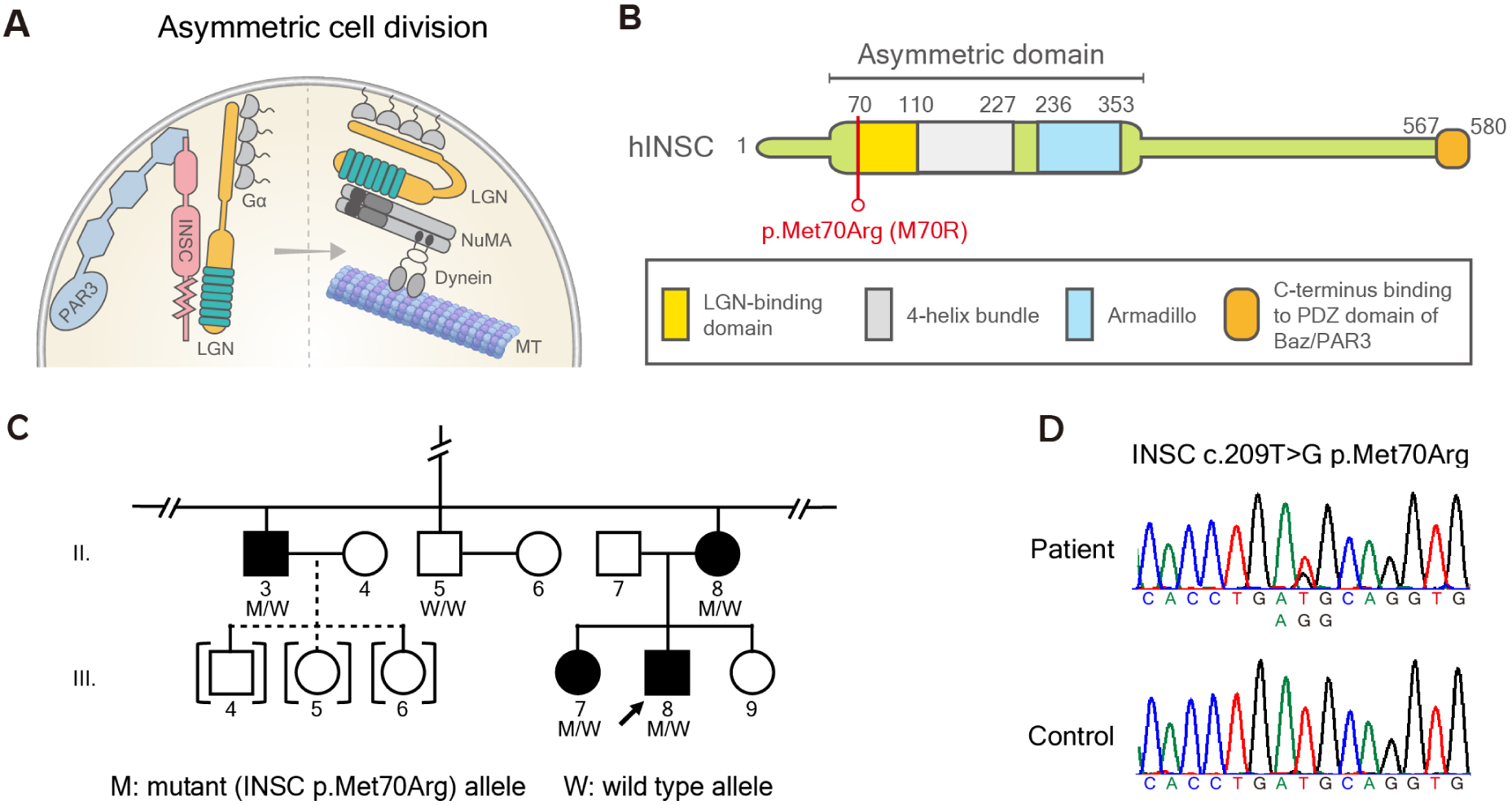
Identification of CMT-associated *INSC^M70R^* variant. **(A)** Schematic figure of ACD in neuroblast. Inscuteable (INSC), an adaptor protein in ACD, regulates microtubules by binding with PAR3 and LGN. **(B)** The *hINSC^M70R^* mutation resides in the LGN-binding domain of the hINSC protein. **(C)** The partial pedigree of a CMT2 family carrying the *INSC* mutation. “M” represents the INSC mutant allele, and “W” stands for wild type allele. Open symbol: unaffected; filled symbol: affected; symbol with a diagonal line: deceased; arrow: proband; squares: males; circles: females; brackets with dashed line: adopted into the pedigree. Data regarding the full pedigree can be accessed by contacting the corresponding authors. **(D)** Sanger sequencing traces confirming the c. 290T>G (p.Met70Arg, M70R) variant.

The *Drosophila* femoral chordotonal organ (FeCO) is the largest proprioceptive organ and comprises over a hundred of mechanosensory neurons rich in MT (*18*). It is considered a functional analogue to the muscle spindle in vertebrates (*19*), playing a critical role in detecting mechanical stretches such as muscle tension and joint position (*20*). The anatomical structure and mechanosensory function of fly FeCO neuron has been intensively investigated. Proprioceptive cell death is known to cause neurological disorders (*21*). Therefore, the fly FeCO neurons provide an excellent platform to unravel the underlying mechanisms of peripheral neuropathy. Here, we describe the discovery of the pathogenic variant p.Met70Arg (M70R) in the *INSC* gene to be associated with Charcot-Marie-Tooth disease type 2 (CMT2). We developed a corresponding fly model to study the effect of the PIL complex in the adult PNS. Our study demonstrated the regulation of MT stability in the adult PNS by the PIL complex and suggests that enhancing MT acetylation as a potential therapeutic strategy for addressing CMT disease caused by the *INSC* mutation.

## Results

### (1) Identification of *INSC* p.Met70Arg in a pedigree with autosomal dominant CMT2

We report a three-generation family with autosomal dominant CMT2, in which eight affected and four unaffected individuals were recruited (Fig.1C). The genetic diagnosis of the proband (III-8) remained unsolved after extensive genetic analysis. All patients presented with slowly progressive distal sensory loss and gait unsteadiness with absent or mild weakness in the limbs with onset ranging from age 7 to 29 years (Table 1, fig.S1). The two eldest patients (II-2 and II-3) had distal weakness after 55 years of age and required a walker or cane to assist with ambulation (movie S1). The nerve conduction studies revealed axonal sensorimotor polyneuropathy with sensory predominant features (table S1). Genome wide linkage analysis in this family mapped the CMT2 locus to a 17-Mb interval on chromosome 11p15.1-15.4, flanked by the markers rs231359 and rs7118901 (Fig.S2). A multipoint LOD score of 3.0 established significant linkage to the genomic interval 11:2694606-19743250 (hg37). Whole genome sequencing of patients III-3 and III-8 (table S2) identified a heterozygous missense variant, c.209T>G (p.Met70Arg), in the *INSC* gene (RefSeq NM_001031853.5) (Fig.1D), which localizes to the linkage interval. The variant fully segregated with the disease and was absent in 1517 healthy controls from the Taiwan biobank database. The p.Met70Arg variant is located within the N terminus of the LBD of INSC, a motif that is functionally conserved across species. Bioinformatic tools predicted the *INSC* p.Met70Arg variant to be a disease-causing by CADD v1.6 (*22*) (Phred score 23.5) and MutationTaster2 (*23*) (probability value: 0.989). Pathogenicity was also supported by the variant being absent in the gnomAD (*24*) and BRAVO databases (https://bravo.sph.umich.edu/freeze8/hg38/), which in total contain genetic variants from over 208 thousand human genomes.

**Table 1.**
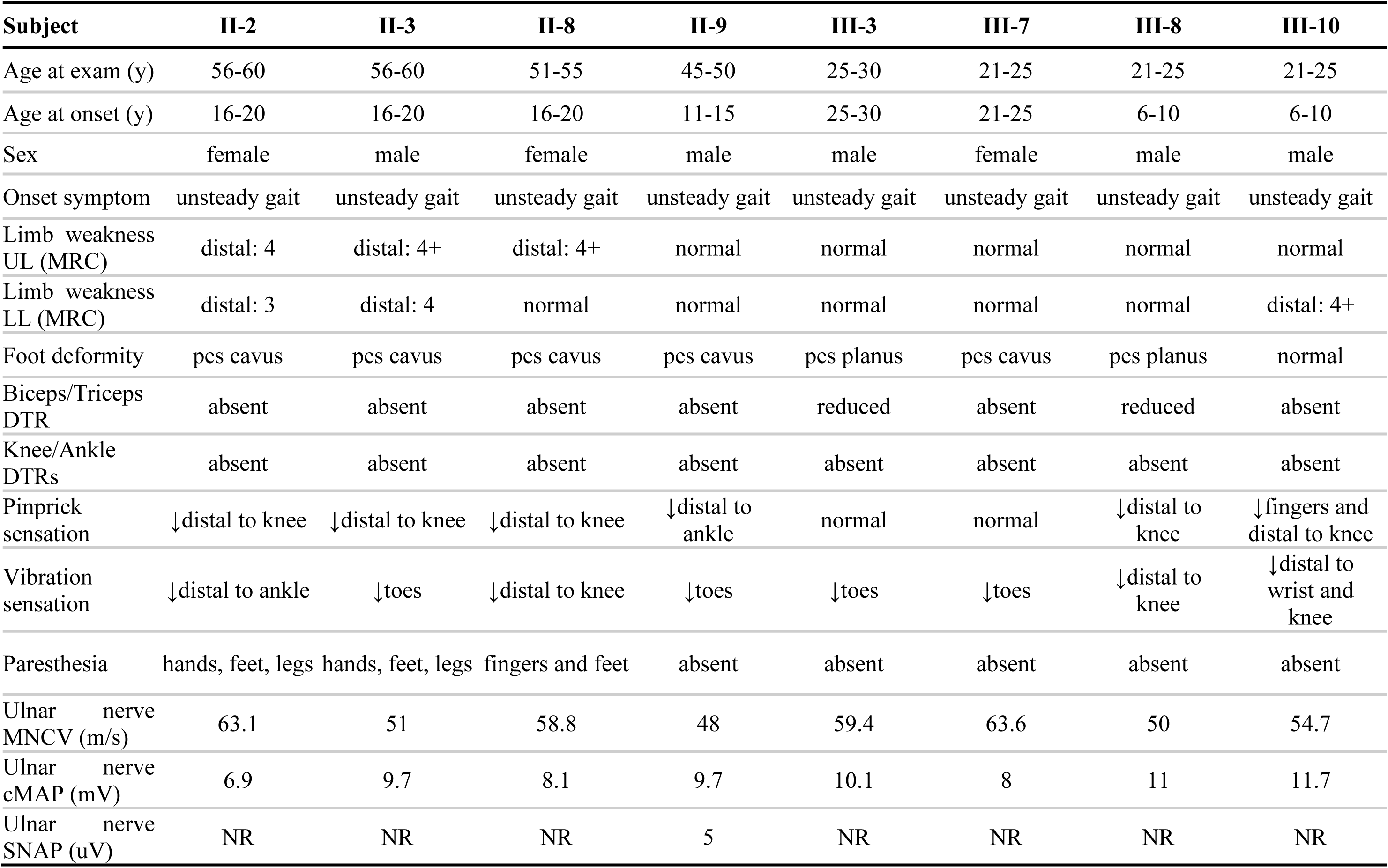
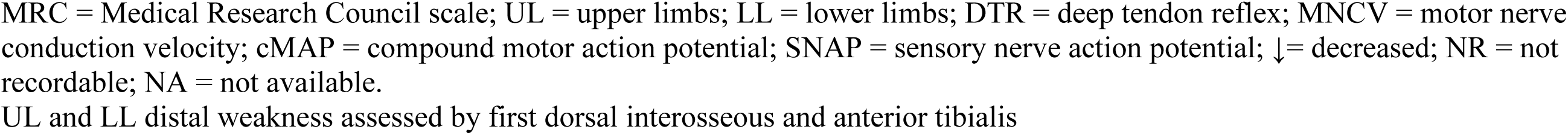
Clinical manifestations of the affected individuals carrying *INSC* p.Met70Arg.

| **Subject** | **II-2** | **II-3** | **II-8** | **II-9** | **III-3** | **III-7** | **III-8** | **III-10** |
| --- | --- | --- | --- | --- | --- | --- | --- | --- |
| Age at exam (y) | 56-60 | 56-60 | 51-55 | 45-50 | 25-30 | 21-25 | 21-25 | 21-25 |
| Age at onset (y) | 16-20 | 16-20 | 16-20 | 11-15 | 25-30 | 21-25 | 6-10 | 6-10 |
| Sex | female | male | female | male | male | female | male | male |
| Onset symptom | unsteady gait | unsteady gait | unsteady gait | unsteady gait | unsteady gait | unsteady gait | unsteady gait | unsteady gait |
| Limb weakness UL (MRC) | distal: 4 | distal: 4+ | distal: 4+ | normal | normal | normal | normal | normal |
| Limb weakness LL (MRC) | distal: 3 | distal: 4 | normal | normal | normal | normal | normal | distal: 4+ |
| Foot deformity | pes cavus | pes cavus | pes cavus | pes cavus | pes planus | pes cavus | pes planus | normal |
| Biceps/Triceps DTR | absent | absent | absent | absent | reduced | absent | reduced | absent |
| Knee/Ankle DTRs | absent | absent | absent | absent | absent | absent | absent | absent |
| Pinprick sensation | ↓distal to knee | ↓distal to knee | ↓distal to knee | ↓distal to ankle | normal | normal | ↓distal to knee | ↓fingers and distal to knee |
| Vibration sensation | ↓distal to ankle | ↓toes | ↓distal to knee | ↓toes | ↓toes | ↓toes | ↓distal to knee | ↓distal to wrist and knee |
| Paresthesia | hands, feet, legs | hands, feet, legs | fingers and feet | absent | absent | absent | absent | absent |
| Ulnar nerve MNCV (m/s) | 63.1 | 51 | 58.8 | 48 | 59.4 | 63.6 | 50 | 54.7 |
| Ulnar nerve cMAP (mV) | 6.9 | 9.7 | 8.1 | 9.7 | 10.1 | 8 | 11 | 11.7 |
| Ulnar nerve SNAP (uV) | NR | NR | NR | 5 | NR | NR | NR | NR |

MRC = Medical Research Council scale; UL = upper limbs; LL = lower limbs; DTR = deep tendon reflex; MNCV = motor nerve conduction velocity; cMAP = compound motor action potential; SNAP = sensory nerve action potential; ↓ = decreased; NR = not recordable; NA = not available.
UL and LL distal weakness assessed by first dorsal interosseous and anterior tibialis

### (2) Loss-of-function of PIL complex of the adult PNS cause proprioceptive defects in aging fly

We first depleted *Drosophila Insc* (*dInsc*) by RNAi to evaluate whether it causes any locomotor defects as seen in human patients. We employed an inducible pan-neuronal *elav-GS-Gal4* driver, which is a modified Gal4/UAS system. This enabled transgene expression upon RU486 drug treatment, thereby allowing adult-onset knockdown of the gene and avoiding any developmental influences. In a locomotor assay, *dInsc-RNAi* flies exhibited normal behavior at day 3 post-eclosion, but showed a climbing deficit at week 1, indicating adult-onset degeneration (Fig.2A). To investigate whether the same machinery utilized to regulate ACD in neuroblasts during brain development is also required in the adult PNS, we knocked down other PIL complex in the adult PNS. Interestingly, knocking down *bazooka* (the fly homolog of human *PAR3*) or *pins* (the fly homolog of human *LGN*) also resulted in climbing defects (Fig.2A). Rescue experiments were performed using the *dInsc^InSITE^* Gal4 driver, an enhancer trap allele with Gal4 insertion in the *dInsc* gene, resulting in the disruption of *dInsc* expression. Heterozygous *dInsc^InSITE^* flies exhibited impaired locomotion that was exacerbated in the *dInsc-RNAi* background, suggesting a dosage-dependent phenotype. The locomotor impairment was rescued by transgenic expression of human *INSC* (*hINSC^WT^*) but not the disease-associated *hINSC^M70R^* (Fig.2B), suggesting that the locomotor function of INSC is conserved between human and flies, and the p.M70R mutation represents a loss-of-function mutation, likely rendering haploinsufficiency.

**Fig. 2.**
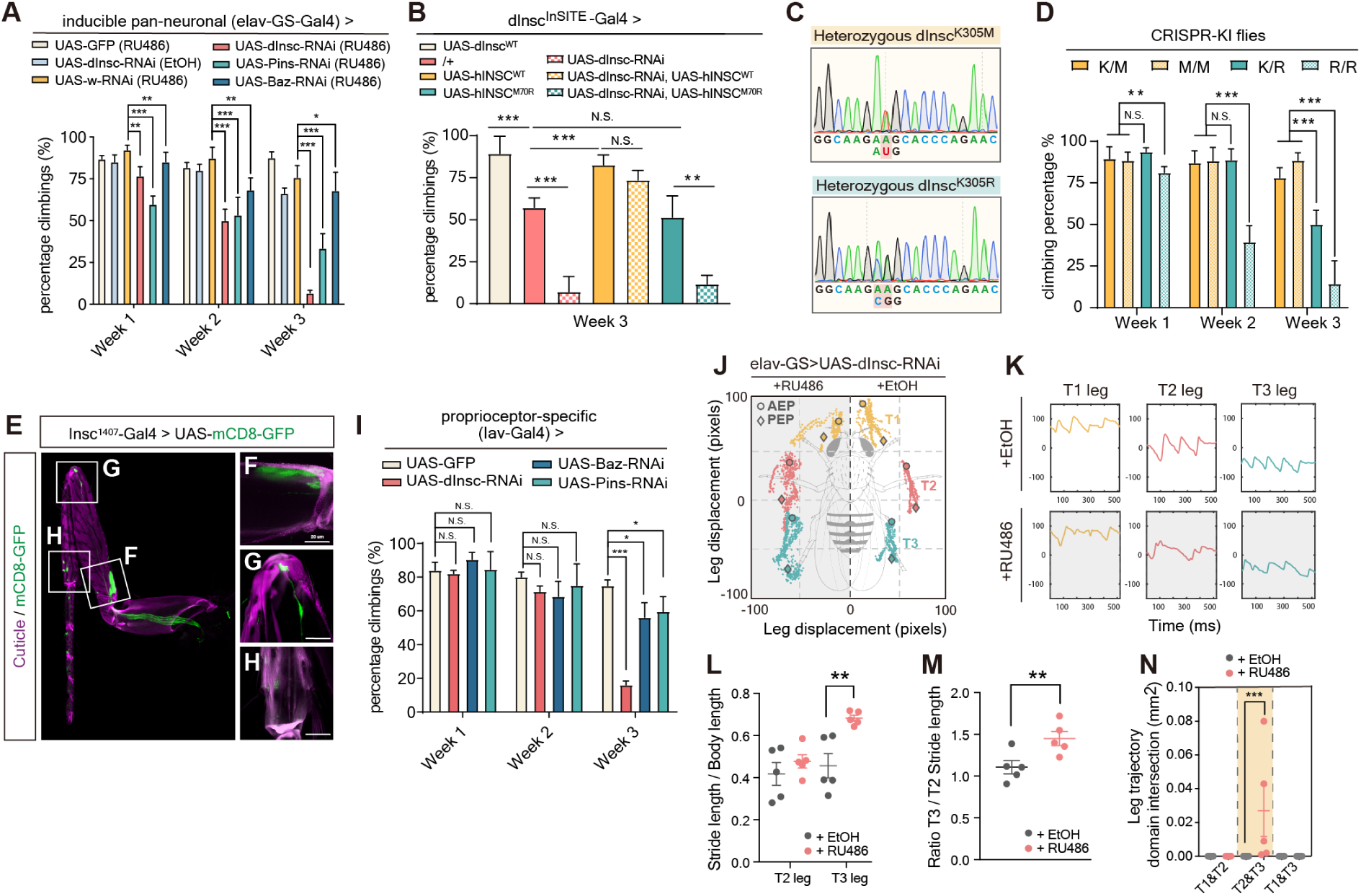
Adult-onset depletion of PIL complex causes locomotor and proprioceptive defects. **(A)** Quantification of the climbing activity of 1– to 3-week-old adult flies with *dInsc*-RNA, *Baz*-RNAi and *Pins*-RNAi under the control of inducible pan-neuronal driver (*elav-GS*-Gal4) upon feeding with RU486 or solvent control (EtOH), comparing with the age-matched controls (*mCD8*-GFP) and (*w*-RNAi). **(B)** Quantification of the climbing activity of 1– and 3-week-old flies with overexpress *dInsc*-WT, /+, *hINSC^W^*, *hINSC^M70R^*, *dInsc*-RNAi and the groups co-overexpress *dInsc*-RNAi with *hINSC^WT^* or *hINSC^M70R^*under the control of *dInsc^InSITE^*-Gal4 driver. **(C)** Sanger sequencing traces confirming the heterozygous *dInsc^K305M^* and *dInsc^K305R^* CRISPR-KI flies. **(D)** Quantification of the climbing activity of 1– to 3-week-old heterozygous + / *dInsc^K305M^*(*K/M*) and + / *dInsc^K305R^* (*K/R*), homozygous *dInsc^K305M^* / *dInsc^K305M^* (*M/M*) and *dInsc^K305R^*/ *dInsc^K305R^* (*R/R*) CRISPR-KI flies. **(E-H)** Thoracic segments 1 (T1) leg of an adult fly expressing *mCD8*-GFP (green) under the control of *dInsc^1407^*-Gal4. Scale bars: 50 µm **(F-H)** Magnified images of the femoral chordotonal organ (F), stretch receptor (G), tibiotarsal chordotonal organ (H, respectively. Scale bars: 20 µm. Magenta is the auto-fluorescence of the cuticle. **(I)** Quantification of the climbing activity of 1– to 3-week-old *dInsc*-RNAi, *Baz-*RNAi and *Pins*-RNAi flies under the control of *Iav*-Gal4, compared with the age-matched controls (*mCD8-GFP*). **(J)** Body-centered leg trajectories plot of 3-week-old *elav-G*S>UAS-*dInsc*-RNAi flies feeding with RU486, compared with the age-matched solvent feeding controls. **(K)** The leg displacement plots of T1 (yellow), T2 (pink) and T3 (cyan) legs from the same groups as (J). **(L)** Quantification of T2 and T3 leg stride length normalized to body length. **(M)** Quantification of the ratio T3/T2 stride length of RU486-fed group, compared with the solvent-fed control. **(N)** Quantification of the leg trajectory domain intersection of RU486-fed group, compared with the solvent-fed control

Given the evolutionary conservation of INSC in neurodevelopment, the impact of human *INSC* mutations underlying peripheral neuropathy was investigated by establishing a fly model. To generate disease-relevant flies with the p.M70R mutation, the corresponding amino acid residue (K305) in the endogenous *dInsc* locus was edited using seamless CRISPR-Cas9 techniques. The location of the amino acid residue (K305) corresponded to position 70 in the human *INSC* gene however the amino acid was not conserved. Two alleles, *dInsc^K305R^*(p.Lys305Arg) to mimic the disease variant and *dInsc^K305M^* (p.Lys305Met) to represent the human wild type were engineered (Fig.2C, fig.S3). We also examined the genetic properties of the two knock-in alleles *dInsc^K305M^* and *dInsc^K305R^*. While *dInsc^K305M^*behaved similarly to the wild type in climbing activity, *dInsc^K305R^*flies exhibited a dosage-dependent, progressive locomotion defect, supporting the notion that *hINSC^M70R^* is a haploinsufficient variant (Fig.2D).

To investigate the post-developmental expression of *dInsc* in the PNS of flies, the *dInsc^1407^-Gal4* driver was employed, which has been extensively utilized in previous studies (*25*, *26*) to specifically drive the expression of *mCD8-GFP* in cells expressing *dInsc*. As previously reported, we confirmed *dInsc* expression in the larval brain and gut during larval development (*7*). Furthermore, our results revealed additional labeling in the leg disc but not the wing disc (fig.S4A). To study the subcellular localization of Insc, we generated a *UAS-dInsc-EGFP* transgene which was co-expressed with *Insc^1407^-Gal4*. The apical localization of *dInsc-EGFP* in a crescent-shaped pattern in the larval neuroblasts was confirmed (fig.S4B) as previously reported in an immunostaining study (*8*). Notably, in the adult leg, the *mCD8-GFP* signal was present in proprioceptive structures, including the femoral chordotonal organ (FeCO), stretch receptors, and tibiotarsal chordotonal organs (tiCO) (Fig.2, E to H). The locomotor impairment resulting from downregulating the PIL complex reflected an impact on proprioception as the knockdown of these genes with the proprioceptive organ-specific (*Iav-Gal4*)-*Gal4* showed similar phenotypes (Fig. 2I). An automated leg tracking system was utilized to analyze and quantify leg trajectory in aged flies. By comparing the gait signature between the control and *dInsc* knockdown flies, significant changes were observed in gait patterns. Specifically, the *dInsc-RNAi* flies exhibited poor footprint regularity (Fig.2, J and K), increased stride length in the hind (T3) leg (Fig.2L) and the ratio of hind/mid (T3/T2) legs (Fig.2M), as well as uncoordinated leg displacement with an enlarged leg intersection domain (Fig.2N). These alterations in gait resembled the walking difficulties and movement dysfunction observed in patients with CMT disease (fig.S5; movie S2 and S3).

### (3) Neurons of loss-of-function of PIL complex displayed adult-onset necrosis

To visualize degenerating neurons, dual-staining with propidium iodide (PI) as a marker for degeneration and DAPI to label the cell nuclei was performed (*27*, *28*). The colocalization of both markers enabled the distinguishment between healthy (DAPI+, PI-), dying (DAPI+, PI+), or dead (DAPI-, PI+) cells (Fig.3A). The loss of *hINSC* in the SH-SY5Y neuroblastoma cells induced necrosis, as 2 independent clones stably expressing *hINSC-shRNA* exhibited a higher ratio of DAPI-PI colocalization compared to the control clone (Fig.3, B and C). In FeCO neurons of PIL knockdown flies, no difference was observed in PI intensity or colocalization of DAPI and PI signals in week 1. However, by week 3, elevated PI staining and increased DAPI-PI colocalization was observed, which was inversely correlated with locomotor decline and reflected the progression of neurodegeneration over time (Fig.3, D and E). The homozygous *dInsc^K305R^* (*R/R*) flies showed higher DAPI-PI colocalization and more severe locomotor defects compared to the heterozygous *dInsc^K305R^* (*K/R*) and wild-type *dInsc* (*K/K*) flies (Fig.3F and fig.S6A). Additionally, the necrosis associated with the functional decline, and the neuronal loss was alleviated by transgenic expression of the human wild-type transgene (Fig.3G, and fig.S6B).

**Fig. 3.**
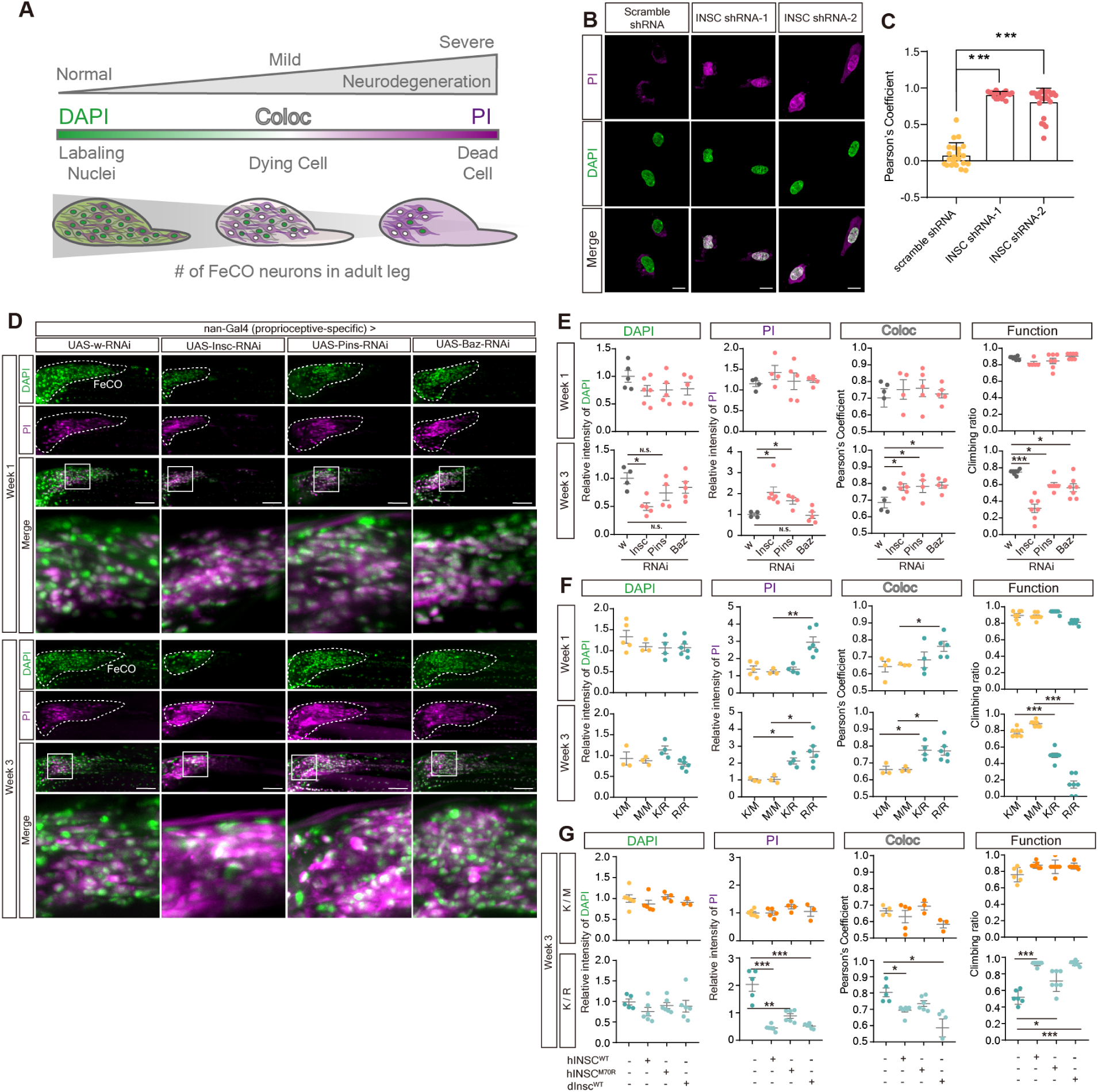
Counter-correlation between necrosis and locomotor activity in PIL loss of function flies. **(A)** Schematic figure analyzing the relative intensity and colocalization of DAPI and PI staining indicated the progression of necrotic cell death *in vivo*. The intensity and colocalization of DAPI and PI may represent the stage of necrosis. For instance, in the healthy neurons, the signal of DAPI is largely stronger than PI; In the situation of progressive necrosis, DAPI may highly colocalize with PI; In the stage of severe neurodegeneration, most of the neuronal cell death, we can just detect a weak signal of DAPI, but a relative stronger signal of PI. **(B)** PI staining in scramble shRNA and *hINSC*-shRNA SH-SY5Y cell. Magenta is PI staining. Green is DAPI staining. Scale bar: 10 µm **(C)** Pearson’s coefficient of colocalization between PI and DAPI in *hINSC*-shRNA-1 and *hINSC*-shRNA-2, compared with scramble shRNA in (B). **(D)** Representative confocal images of *dInsc*-RNAi, *Pins*-RNAi and *Baz*-RNAi co-staining with PI (magenta) and DAPI (green) in FeCO neurons of 1– and 3-week-old flies, compared with *w*-RNAi control. The FeCO neurons are encircled by the dashed lines. Scale bar: 5 µm **(E)** Quantification of the relative intensity and colocalization (coloc) of DAPI and PI in (D), and comparing the results with the functional assay. The relative intensity is normalized with *w*-RNAi. **(F)** Quantification of the relative intensity and colocalization of DAPI and PI in (fig.S6A), and comparing the results with the functional assay. The relative intensity is normalized with *W^1118^*(K/K) flies. **(G)** Quantification of the relative intensity and colocalization of DAPI and PI in (fig.S6B), and comparing the results with the functional assay. The relative intensity is normalized with *K/M* and *K/R* CRISPR-KI flies, respectively.

### (4) p.M70R variant reduced Insc expression in old age and differentially affects interaction with LGN and PAR3

We collected cDNA samples from the blood of both affected and unaffected family members and revealed a significant decrease in *hINSC* mRNA levels in young affected individuals compared to young healthy individuals. The *hINSC* mRNA abundance further decreased in older affected individuals (Fig.4A). Similarly, in flies, we observed an age-dependent decline in hINSC^M70R^ protein levels, particularly in the dendrites of FeCO neurons. This observation suggests that the reduction in protein expression is inherent to the p.M70R mutation (Fig.4, B and C). Since the p.M70R mutation resides within the LGN-binding motif of hINSC, the effect of the mutation on the association with the PIL complex was investigated. In human SH-SY5Y cells, co-immunostaining of hINSC with LGN showed decreased hINSC-LGN colocalization in hINSC^M70R^-expressing cells (Fig.4, D and E). Conversely, co-immunostaining with HA-PAR3 showed higher colocalization between PAR3 and hINSC^M70R^ (Fig.4, F and G). Similarly, confocal imaging of FeCO neurons in 3-week-old flies revealed an increase in colocalized puncta of *hINSC^M70R^-EGFP* and *Bazooka-mCherry*, and a decreased co-localization with *Pins-mCherry* compared with *hINSC^WT^*control (Fig.4, H to J). These findings highlight the impact of the mutation on mRNA abundance, protein level, and LGN binding.

**Fig. 4.**
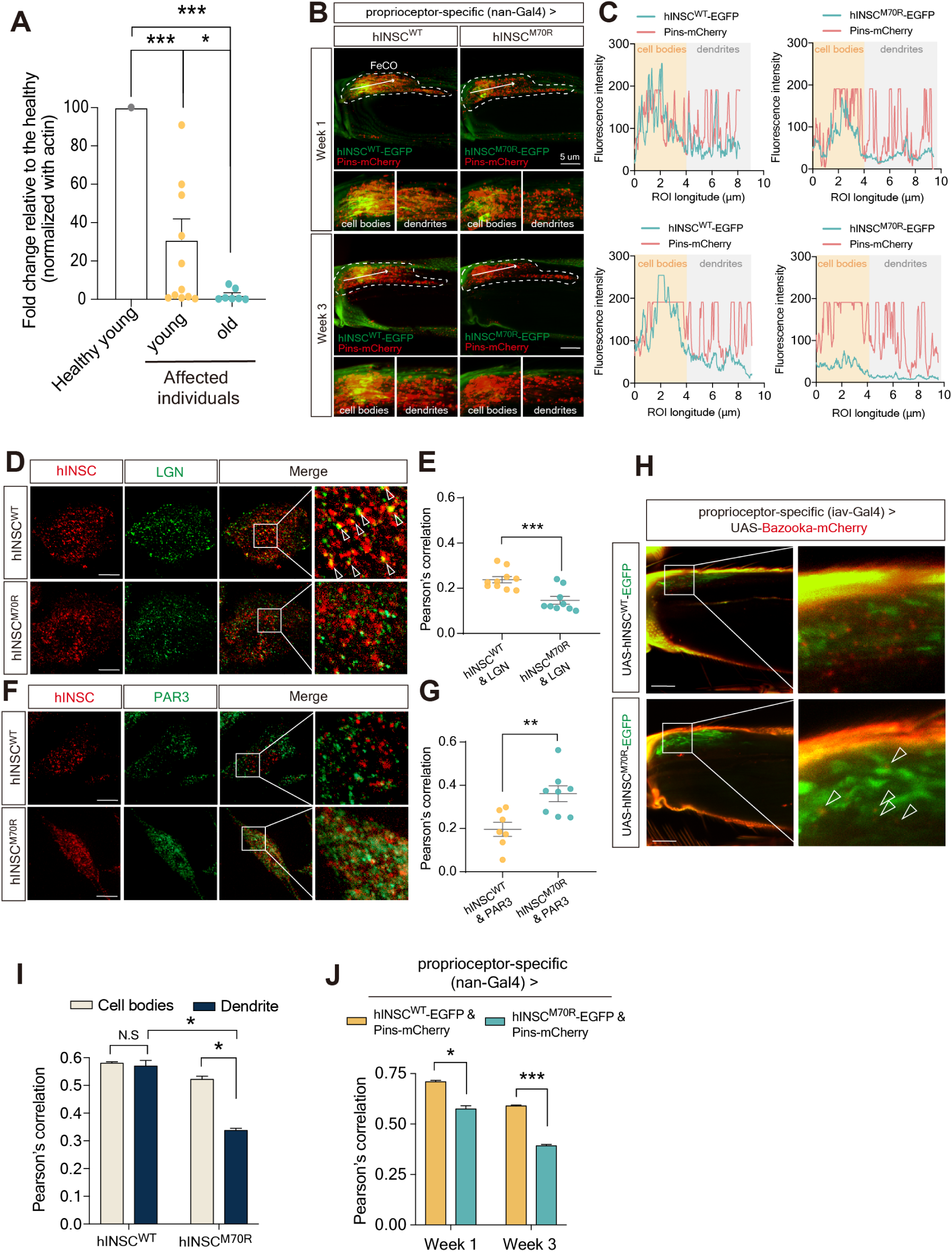
hINSC-M70R protein exhibited decreased levels and altered association with PIL complex. **(A)** Relative mRNA abundance of *hINSC* in young and old affected individuals, compared with healthy young control. By RT-PCR the mRNA abundance of INSC from the whole blood of the patients. Compared to the healthy young control. **(B)** Representative images of co-expressing *hINSC^M70R^-EGFP* (green) and *Pins*-mCherry (red) in 1– and 3-week-old flies under FeCO-expressing *nan*-Gal4, compared with the age-matched controls (*hINSC^WT^-EGFP*). The white arrow indicates the direction of the fluorescence intensity profile. Scale bars: 5 µm. Shown below are the magnified insets of cell bodies and dendrites in FeCO neurons, respectively. Scale bars: 5 µm **(C)** Fluorescence intensity profiles were generated to visualize co-localization of *hINSC^M70R^-EGFP* and *Pins-mCherry* of cell bodies and dendrites in 1– and 3-week-old flies, compared with the age-matched controls (*hINSC^WT^-EGFP*). The linear region of interest (ROI) was drawn manually from left to right. **(D)** Immunostaining of SH-SY5Y cells transfected with FLAG-hINSC^M70R^ (red) to visualize the colocalization with MYC-LGN (green), comparing with FLAG-hINSC^WT^ (red) control. Scale bar: 10 µm **(E)** Pearson’s coefficient of colocalization of anti-FLAG (red) and anti-MYC (green) fluorescence in (D). **(F)** Immunostaining of SH-SY5Y cells transfected with FLAG-hINSC^M70R^ (red) to visualize the colocalization with HA-PAR3 (green), comparing with FLAG-hINSC^WT^ (red) control. Scale bar: 10 µm **(G)** Pearson’s coefficient of colocalization of anti-FLAG (red) and anti-HA (green) fluorescence in (F). **(H)** Representative images of co-expressing *hINSC^M70R^-EGFP* (green) and *Baz-mCherry* (red) in 1– and 3-week-old flies under *Iav*-Gal4, compared with the age-matched controls (*hINSC^WT^-EGFP*). Scale bars: 5 µm **(I)** Pearson’s coefficient of colocalization *hINSC^WT^* and *hINSC^M70R^* with *Pins* in (B) in cell bodies and dendrite in FeCO neurons of 1-week-old flies. **(J)** Pearson’s coefficient of colocalization of whole FeCO neurons of 1– and 3-week-old flies in (B).

### (5) Dysfunction of PIL complex caused tubulin aggregation in adult proprioceptive organs

Our results suggest reduced association of hINSC^M70R^ with LGN may lead to MT instability in the adult PNS, as hINSC competes with NuMA for LGN-binding to regulate MT arrangement during spindle assembly in neuroblasts (*15*). After examining tubulin in the scolopidium, a MT-rich region in adult FeCO neurons, large tubulin puncta were observed to accumulate extracellularly near *dInsc-RNAi* neurons between the muscle fibers (Fig.5, A to C, and fig.S7, A and B). The *hINSC^M70R^* also caused similar aggregates that were absent in the *hINSC^WT^*animals. Likewise, *pins-RNAi*, *bazooka-RNAi*, and the disease-relevant heterozygous *dInsc^K/R^* flies all exhibited tubulin aggregation in the proprioceptive structure, in which the severity of aggregation progressed over time (Fig.5, D and E, and fig.S8, A and B).

**Fig. 5.**
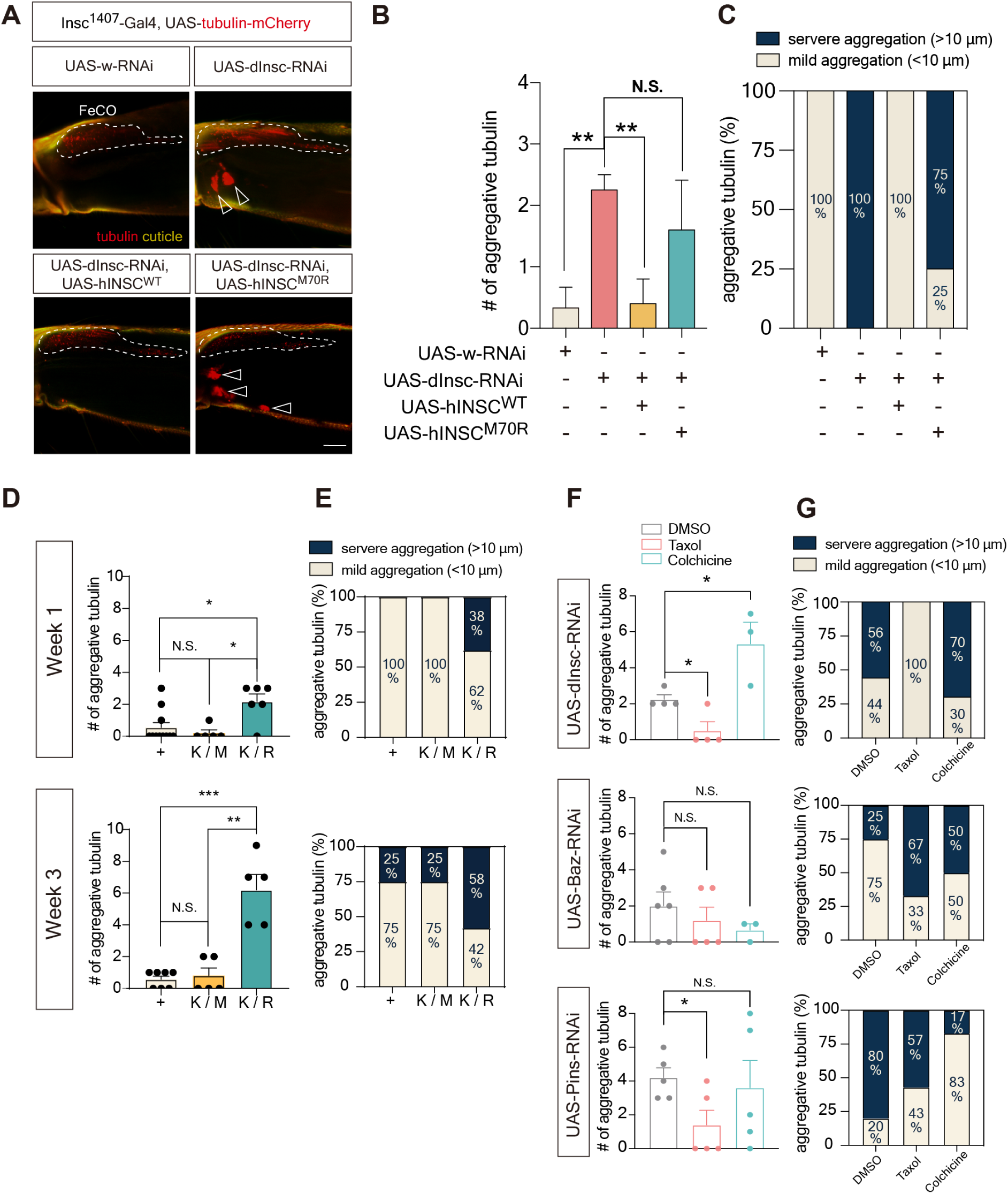
Aging flies of PIL loss-of-function exhibited tubulin aggregation in FeCO neurons. **(A)** Representative confocal images of tubulin aggregation (Red indicates *tubulin*-*mCherry*) of the femur in 3-week-old *dInsc*-RNAi flies under *dInsc^1407^*-Gal4 compared with control (*w*-RNAi), rescued by *hINSC^WT^* but not *hINSC^M70R^*. Yellow indicated the auto-fluorescence of cuticles. The FeCO neurons are encircled by the dashed line. Arrowheads indicate the aggregative tubulin. Scale bars: 5 µm **(B)** Quantification of the number of aggregative tubulins in (A). **(C)** Relative abundance of aggregative tubulins of different sizes in (A). **(D)** Quantification of the number of aggregative tubulins in (fig.S8A). **(E)** Relative abundance of aggregative tubulins of different sizes in (fig.S8A). **(F)** Quantification of the number of aggregative tubulins in (fig.S8B). **(G)** Relative abundance of aggregative tubulins of different sizes in (fig.S8B).

### (6) Defects of PIL complex dysfunction were alleviated by Taxol treatment

As a proof-of-concept experiment, we explored the therapeutic potential of the readily-available MT-stabilization agent Paclitaxel (Taxol), which does not cross the blood-brain barrier (BBB) and can specifically target the PNS without affecting the brain. Although Taxol is known to cause peripheral neuropathy in 60% of patients receiving paclitaxel chemotherapy(*29*), we found that treatment of Taxol at low concentration in the PIL complex knockdown flies reduced the number and size of tubulin accumulations in the proprioceptive structure (Fig.5, F and G, and Fig.6, D to F). Conversely, Colchicine, a MT-destabilizing drug, exerted the opposite effect and further exacerbated the aggregation of tubulin.

**Fig. 6.**
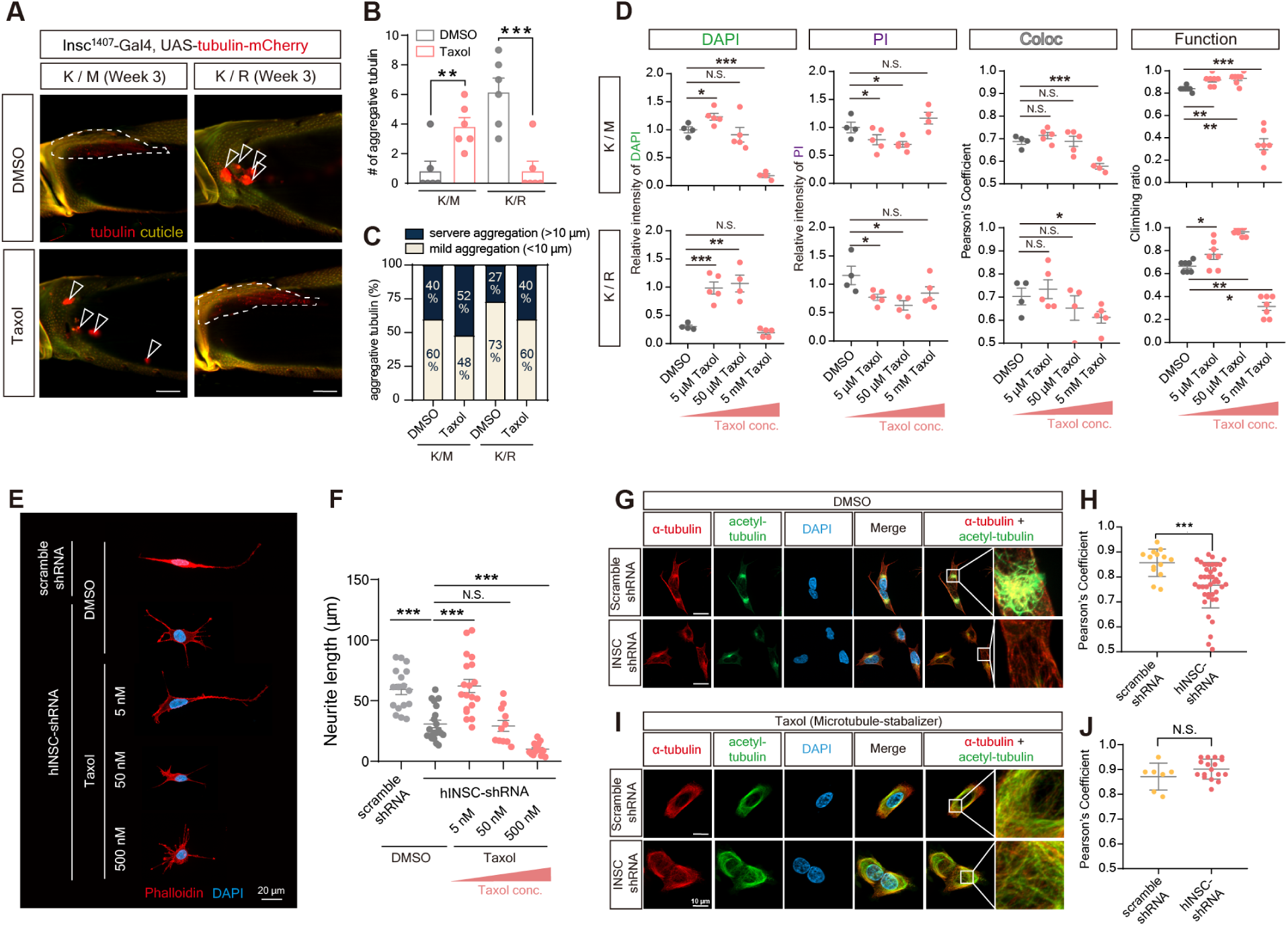
Treatment of optimal concentration of Taxol rescued the morphological and functional defects in cell and aging fly. **(A)** Representative confocal images of tubulin aggregation (Red indicates *tubulin*-*mCherry*) of the femur in 3-week-old with the background of heterozygous *K/M* and *K/R* CRISPR-KI flies under *dInsc^1407^*-Gal4. Flies were treated with microtubule stabilizer Taxol for 7 days, compared with vehicle control (DMSO). Yellow indicated the auto-fluorescence of cuticles. The FeCO neurons are encircled by the dashed line. Arrowheads indicate the aggregative tubulin. Scale bars: 5 µm **(B)** Quantification of the number of aggregative tubulins in (A). **(C)** Relative abundance of aggregative tubulins of different sizes in (A). **(D)** Quantification of the relative intensity and colocalization of DAPI and PI in (fig.S8C), and comparing the results with the functional assay. The relative intensity is normalized with the vehicle control (DMSO) group. **(E)** Immunostaining of *hINSC*-shRNA transfected SH-SY5Y cells and treated with DMSO and Taxol to visualize the neurite length compared with scramble-shRNA control. Red is phalloidin. Blue is DAPI staining. Scale bar: 20 µm **(F)** Quantification of the mean neurite length in (E). **(G)** Immunostaining of hINSC-shRNA transfected SH-SY5Y cells and treated with DMSO (a vehicle control of Taxol) to visualize the colocalization of α-tubulin (red) and acetylated-tubulin (green) compared with scramble-shRNA control. Blue is DAPI staining. Scale bars: 10 µm **(H)** Pearson’s coefficient of colocalization of anti-α-tubulin (red) and anti-acetylated tubulin (green) fluorescence in (G). **(I)** Immunostaining of hINSC-shRNA transfected SH-SY5Y cells and treated with microtubule-stabilizer Taxol to visualize the colocalization of α-tubulin (red) and acetylated-tubulin (green) compared with scramble shRNA control. Blue is DAPI staining. Scale bars: 10 µm **(J)** Pearson’s coefficient of colocalization of anti-α-tubulin (red) and anti-acetylated tubulin (green) fluorescence in (I).

Collectively, these findings suggest that microtubule destabilization resulting from PIL complex dysfunction is a likely cause of PNS degeneration. Consistently, Taxol alleviated tubulin aggregation in the disease-mimicking *dInsc^K/R^* flies compared with *dInsc^K/R^* flies treated with DMSO or the heterozygous *dInsc^K/M^* flies (Fig.6, A to C).

Although Taxol has been proposed to induce Chemotherapy-induced peripheral neuropathy (CIPN) in cancer patients, it has also been reported to increase the neurite length (*30*). We noticed that Taxol induced tubulin aggregation in the *dInsc^K/M^* flies (Fig.6, A to C), suggesting the dose of Taxol is important. To examine whether these distinct effects of Taxol are due to different concentrations, we assessed the dosage effect of Taxol on heterozygous *dInsc^K/R^*flies. We found that low concentrations of Taxol were neuroprotective in alleviating the necrotic status in FeCO neuron and restoring the locomotor activity, while high concentrations exerted toxic effects (Fig.6D, and fig.S8C). In addition, Taxol treatment also ameliorated the shortened neurite length in SH-SY5Y cells with *hINSC-shRNA* (Fig.6, E and F). To examine microtubule stabilization, we immunostained acetylated α-tubulin as a marker and found a decrease in acetylated tubulin in SH-SY5Y cells with *hINSC-shRNA* (Fig.6, G and H). Taxol treatment eliminated the effect of *hINSC-shRNA* on tubulin acetylation (Fig.6, I and J). Together, our data suggest that low dose of Taxol can enhance microtubule stability to rescue the neuronal necrosis and locomotive defect caused by PIL complex dysfunction.

## Discussion

We report here the identification of a large family with a new CMT mutation affecting INSC, a gene known for its role in ACD during neurodevelopment. Our research findings provide evidence that the loss-of-function of PIL complex results in progressive locomotor impairments and abnormal gait in aging flies, which closely resemble the phenotypic characteristics observed in individuals with the p.Mer70Arg mutation. Our investigations further revealed that the PIL complex are required for the control of gait and climbing by stabilizing MT within the adult PNS. Molecularly, haploinsufficiency of PIL complex induces tubulin aggregation, which is likely a remnant of necrotic cells that had disrupted plasma membrane integrity and underwent microtubule destabilization.

Additionally, we demonstrated that treatment with optimal doses of Taxol, a drug known for stabilizing MT, can effectively alleviate the morphological and functional abnormalities associated with the loss of PIL complex function in the adult PNS in Drosophila carrying the same human mutation.

In this study, a combination of genome-wide linkage analysis and whole genome sequencing identified a missense genetic variant in the *INSC* gene, p.Met70Arg, which is associated with dominant CMT2 in this large pedigree. Notably, the *INSC* p.Met70Arg mutation was absent in the gnomAD and BRAVO databases (approximately 273 thousand exomes or genomes), as well as in 1,517 ethnically matched control genomes. Interestingly, whereas CMT patients typically exhibit prominent motor symptoms, the affected individuals in this particular family showed sensory symptoms in the early disease stages, with significant motor symptoms appearing later in life, typically after the age of 50. This distinctive pattern of symptom manifestation raises further interest in the genetic basis of the condition. This intriguing parallel between the fly model and the family’s condition adds weight to our conclusion that the *INSC* p.Met70Arg is indeed the genetic variant responsible for CMT in this specific family.

Modulating MT stability has been shown as a potential target for treating neurodegeneration (*31*). MT destabilization, the core phenotype observed in our study, have been implicated in the pathogenesis of neurodegenerative disorders such as Alzheimer’s disease (AD), Parkinson’s disease (PD), and Amyotrophic Lateral Sclerosis (ALS) (*31*). AD is characterized by the accumulation of neurofibrillary tangles (NFTs) primarily composed of modified Tau protein. Tau protein acts as a stabilizer of MTs, which is vital for axon outgrowth (*32*). Consequently, the loss of tau can lead to decreased MT stability, a condition that can potentially be addressed by treatment with an MT stabilizing agent such as Epothilone D (*33*). Epothilone is reported to enhance axonal growth in CNS injuries and cross the blood-brain barrier (*34*). To explore the therapeutic potential for PNS neuropathy, we utilized Taxol, another FDA-approved MT-stabilizing drug which exhibits low capacity to access the brain (*35*, *36*). Taxol, when administrated at high dose, has been observed to accumulate in the dorsal root ganglia (*37*). Interestingly, we found that a relatively low concentration of Taxol increased neurite length and prevented necrotic cell death, whereas a relatively high concentration had the opposite effect. Consistent with our findings, previous studies have shown that high concentrations of Taxol may induce apoptosis-independent axonal degeneration and aberrant microtubule aggregates in both vertebrate and invertebrate organisms (*38*, *39*). Conversely, low doses of Taxol promote axon regeneration by stimulating axonal growth (*30*). Molecular investigations have demonstrated that high concentrations of Taxol induce the formation of microtubule bundles and increase polymer mass in cells (*40*), whereas lower concentrations merely suppress microtubule dynamics without affecting polymer mass (*30*, *41*).

We have shown the expression of PIL complex in adult PNS mechanosensory neurons, particularly the FeCO neurons. At the molecular level, the regulation of tubulin acetylation involves α-tubulin acetyltransferase 1 (α-TAT1) and histone deacetylase 6 (HDAC6). α-TAT1 plays a critical role in the mechanosensation of the PNS in both Drosophila and mammals by modulating α-tubulin acetylation. Loss-of-function mutations in α-TAT1 have been observed to result in reduced mechanosensitivity in affected individuals (*42*, *43*). Furthermore, disrupted MT stability can lead to abnormal tubulin distribution. For example, the absence of Spastin, a regulator of MT stability, can lead to neurite swelling (*44*). Moreover, our investigation revealed a reduction in tubulin acetylation upon PIL loss of function due to haploinsufficiency of the *INSC* gene. Previous studies have established that tubulin acetylation serves as a marker of stable MT in neuronal cytoskeletons, and its decrease can contribute to the impairment of MT-mediated axonal transport, potentially leading to neurodegenerative disorders like CMT disease (*45*). Importantly, this abnormality can be remedied by promoting the acetylation of α-tubulin (*46*).

In conclusion, our study presents compelling clinical and experimental evidence establishing a causal relationship between PIL haploinsufficiency and adult-onset hereditary neuropathy. Our findings underscore the significance of the PIL complex in regulating microtubule stability for adult proprioceptive function. Leveraging the robust genetic tools available in Drosophila, our model has allowed for multiple genetic manipulations and holds promise as a valuable resource for investigating therapeutic interventions. Through the integration of clinical observations and genetic models, our study elucidates the molecular mechanisms underlying necrotic neurodegeneration associated with PIL complex. Based on our findings, we propose enhancing microtubule acetylation as a potential therapeutic strategy for addressing CMT2 associated with mutations in PIL complex.

## Data Availability

All data are available in the manuscript or the supplementary material. All fly strains, cell lines, and DNA vectors are readily available through the corresponding author. Antibodies are commercially available.

## Acknowledgments

We are grateful for the patients who consented to being a part of this study. We thank all members of the Chan and Lee labs for critical comments on this manuscript. We thank the Blooming Drosophila Stock Center, Vienna Drosophila RNAi Center, and Developmental Studies Hybridoma Bank for reagents, WellGenetics Inc. for the generation of transgenic flies, and the Biomedical Resource Core and the imaging core at the First Core Labs in National Taiwan University College of Medicine for technical support in molecular cloning, image acquisition and analysis.

## Funding

This work was supported by grants from the National Science and Technology Council of Taiwan (111-2320-B-002-049-MY3 and 112-2314-B-002-016 to C.-C.C;109-2314-B-075-044-MY3 and 112-2314-B-075-034-MY3 to Y.-C.L), National Health Research Institutes (EX112-11228NI to C.-C.C), National Taiwan University (112L895403), and Brain Research Center, National Yang Ming Chiao Tung University from The Featured Areas Research Center Program within the framework of the Higher Education Sprout Project by the Ministry of Education (MOE) in Taiwan.

## Author contributions

Conceptualization: JYY, YCL, CCC

Methodology: JYY, YCH, FYT

Investigation: JYY, HCC, CTH, CTC, YST, YCL, SYH, MK, YCL, CCC

Visualization: JYY

Supervision: YCL, CCC

Writing—original draft: JYY, YCL, CCC

Writing—review & editing: JYY, SYH, MK, YCL, CCC

## Competing interests

The authors declare no competing interests.

## Supplementary Materials

Materials and Methods

Figs. S1 to S8

Tables S1 to S7

Movies S1 to S3

References (47 – 54

## Conclusion

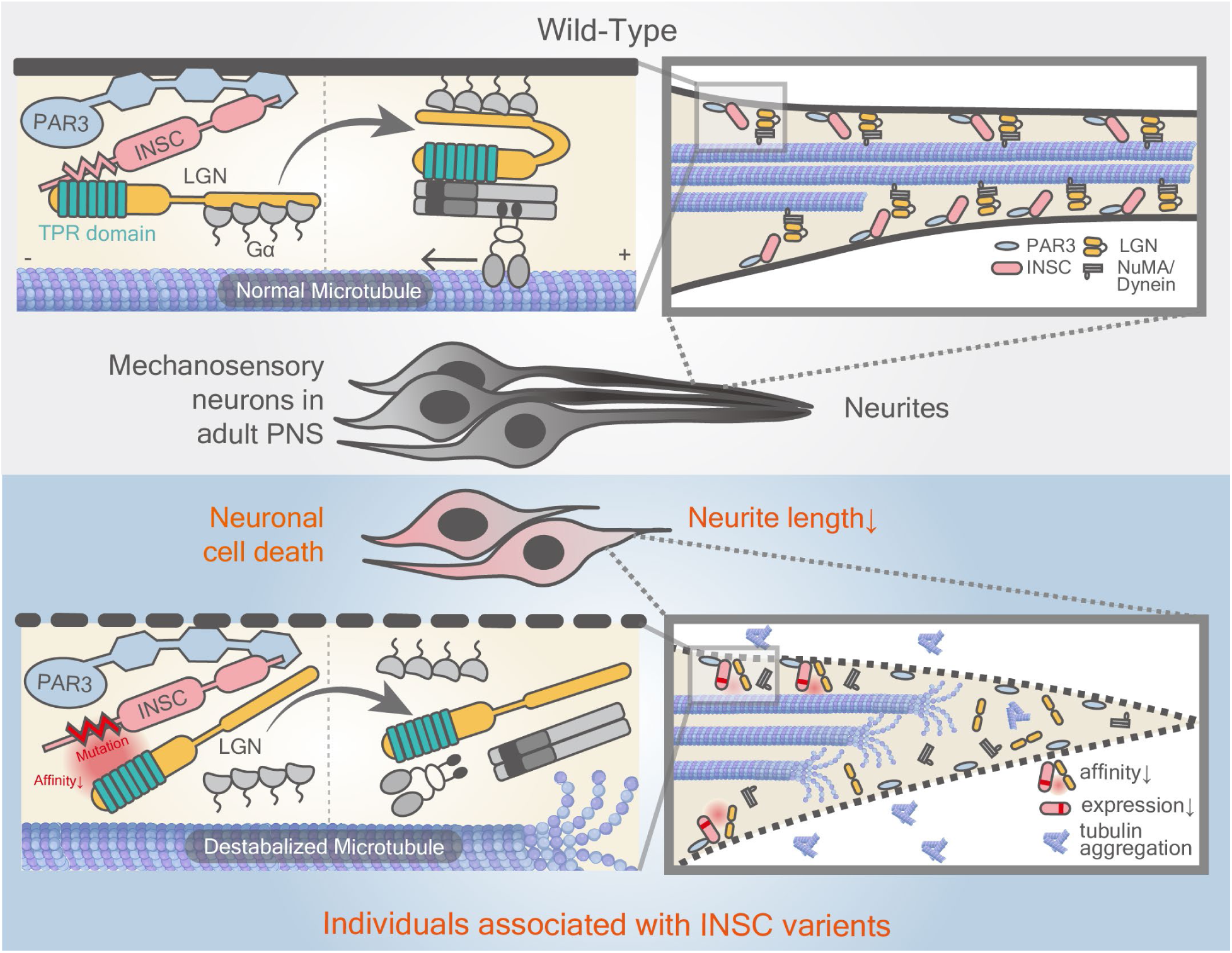
The possible role of PIL complex for microtubule maintenance and neurodegeneration in PNS. Under physiological conditions, PIL complex maintain the mechanosensory neurons in adult PNS by regulating microtubule stability. In CMT2 patients associated with *INSC* mutations, decreased association between INSC and LGN causes microtubule destabilization, resulting in neurite length shortening, neuronal cell death, and functional decline

## Notes

### Competing Interest Statement

The authors have declared no competing interest.

### Author Declarations

the study was approved by the Institutional Review Board of Taipei Veterans General Hospital, Taiwan (TPEVGH IRB No.:2020-02-016B).

