## Supplemental file for "Disruption of the PAR3/INSC/LGN complex causes microtubule instability and peripheral neuropathy"

Science
AAAS

Supplementary Materials for

### Disruption of the PAR3/INSC/LGN complex causes microtubule instability and peripheral neuropathy

Jui-Yu Yeh *et al.*

**The PDF file includes:**

Materials and Methods
Figs. S1 to S8
Tables S1 to S7
References 47 - 54

**Other Supplementary Materials for this manuscript include the following:**

Movies S1 to S3

#### Materials and Methods

##### Subjects

Twelve members of the families with molecularly unassigned autosomal-dominantly CMT2 were enrolled into the study, including 8 patients and 4 unaffected individuals (Fig.1C). Mutations in 124 genes associated with inherited neuropathies were excluded in the proband (III-10) by targeted resequencing. Nerve conduction studies were performed by standard techniques utilizing a Medelec MS25 electromyograph (Mistro, Surrey, U.K.) with surface electrode stimulations and recordings. Written informed consents were obtained from all subjects and the study was approved by the Institutional Review Board of Taipei Veterans General Hospital, Taiwan (TPEVGH IRB No.:2020-02-016B).

##### Genetics analyses

Whole genome sequencing was performed in the two patients (III-8 and III-10) from the CMT2 family. Illumina HiSeq2500 platform with a paired-end 2 x100 bp protocol was used for sequencing. Because of the autosomal dominant inheritance, only heterozygous variants shared by the two patients were taken for further analyses. Variants present in the 1517 unrelated healthy Taiwanese genomes from the Taiwan biobank (https://taiwanview.twbiobank.org.tw/) and those that do not alter coding sequences were excluded. Sanger sequencing was then performed to determine whether any of the remaining variants completely co-segregated with CMT2 dHMN in the family (table S1). Mutation Taster (*47*) and Combined Annotation Dependent Depletion (CADD) (*48*) were used to predict the pathogenicity of the identified variants. For linkage analysis, the twelve individuals in the family with CMT2 were genotyped with ~560K markers by the Illumina Infinium assay using the Infinium CoreExome-24 v1.3 BeadChip (Illumina, San Diego, CA). Multipoint linkage was performed using the MERLIN program.

##### Fly husbandry and stocks

Drosophila stocks and crosses were maintained at 25°C on standard medium following standard fly husbandry. Information on individual fly strains and genotypes for experiments are listed in table S3 and S4 and can be found on FlyBase.

##### Generation of point mutation CRISPR-KI flies

Fly $dInsc^{K305M}$ and $dInsc^{K305R}$ knock-in point mutation were generated using the CRISPR/Cas9 system. The gRNA for CRISPR/Cas9 target sites was designed using UCSC genome browser (https://genome.ucsc.edu/), and ligated into pBFv-U6.2 (Addgene #138400) plasmid.

To clone the donor DNA plasmid, both upstream and downstream homology arms of *dInsc* were amplified from fly genomic DNA by Phusion® High-Fidelity DNA Polymerase (New England Biolab #M0530S) under optimized conditions. The two homology arms were then cloned into pScarlessHD-DsRed (Addgene #64703) plasmid by SOEing PCR, followed by standard heat-shock transformation, colony PCR selection, and Sanger sequencing. Embryo microinjection service was provided by Wellgenetics, Taiwan, to inject the gRNA and donor DNA plasmids into fly embryos with Cas9 activity in the germ cell. Successful integration events were selected by following 3xP3-ScarlessDsRed marker in the progeny, and verified by junctional PCR. After selection, the 3xP3-ScarlessDsRed cassette in the genome of the CRISPR-modified flies was removed by PiggyBac transposase (BDSC #32174). Single fly genomic PCR was performed to verify the CRISPR-KI alleles of *dInsc*. The CRISPR-KI flies were confirmed by Sanger sequencing and background mutations were cleaned by outcrossing to wild-type flies before conducting further morphological and functional tests.

**Generation of transgenic flies**

For the generation of *UAS-dInsc-EGFP*, *UAS-hINSC-WT-FLAG*, *UAS-hINSC-M70R-EGFP*, *UAS-dPins-mCherry*, *UAS-hINSC-WT-EGFP* and *UAS-hINSC-M70R-EGFP*. The transgenesis, from molecular cloning, embryo injections, to screening and balancing of transformants was performed following standard protocol. For cloning, the plasmid, primers and restriction enzymes are listed in table S5 and S6.

**Dissection & fixation & mounting of fly tissue**

For dissection, all tissues from larvae or adult fly were dissected according to protocols as previously described (*49*, *50*).
After dissection, the adult leg was fixed in 4% paraformaldehyde (PFA) for 1 day, while all other tissues were fixed for 20 minutes. The fixants were washed in PBS with 0.4% Triton X-100 (0.4% PBST) three times for ten minutes. The samples were mounted in VECTASHIELD (Vector Laboratories #H-1000) for further analysis.

**immunohistochemistry of fly tissues**

For immunostaining, the fly tissues were blocked in 0.4% PBST with 1% bovine serum albumin (BSA, BioShop Canada Inc. #ALB001) at room temperature for 1 hr, and then washed with 0.4% PBST for 3 times for 10 minutes. After blocking, all tissues were incubated in primary antibody at 4°C for 1 day, washed in 0.4% PBST three times for 10 minutes, followed by secondary antibody incubation at 4°C overnight. The catalog number and the working concentration of both primary and secondary antibodies were described in table S7.

**fluorescent microscopy and confocal imaging**

All the fly tissues were scanned on a Zeiss LSM780 confocal microscope and the SH-SY5Y cell on a Zeiss ApoTome.2 microscope. The acquired images were then analyzed with Zen (Zeiss) software.

**Pharmacological induction and reduction in adult flies**

For the induction of *GeneSwitch-Gal4*, 70 µl of 200 µM RU486 (mifepristone #84371-65-3) in 99% ethanol was added to the food. To bypass development, flies were incubated in normal food before eclosion. Freshly eclosed flies were transferred into the foods that contain RU486 for 7 days, and transferred to a new vial every 3 days.
For regulating microtubules, Taxol or Colchicine was added to the food, respectively. To stabilize microtubules in adult flies, the microtubule-stabilizing drug Taxol (Sigma Aldrich #T1912) in DMSO was added to the food; To destabilize microtubules, 5 µg/ml microtubule-depolymerizing agent Colchicine (Sigma Aldrich #C9754) was added to the food. Both drugs were added for 7 days, and the flies were transferred to a new vial with drug-containing food every 3 days.

**Functional analysis of adult flies**

**Climbing assay for locomotor defect**

The climbing (negative geotaxis) assay examined the percentage of the flies that tapped down to the bottom of the vial to climb up and reach the 8-cm mark on the vial in 10 secs (*51*). At the time of the experiment, groups of flies with specified genotypes were placed in a set of plastic vials. The whole experiment, performed with all the groups in 10 repeats, was recorded by video for further analysis.

**Gait analysis and quantification of walking parameters in adult flies**

The gait analysis by leg tracking was performed following (*52*) with modifications. The setup consists of the 1.5 mm thick behavior chamber between two slides with the diffuser and LED light source on the top side and the high-speed camera (FLIR Blackflys$^{@}$ #BFS-U3-04S2M-CS) with 25 mm camera lens (Computar$^{@}$ #M2518-MPW2) recording on the bottom. Before the experiment, the flies were anesthetized by ice, moved to the 10 mm x 10 mm chamber individually, and kept at room temperature for more than 15 mins for refreshing. After awakening, the walking flies were recording frame-by-frame with 500 fps for leg segmentation and tracking. The leg displacement plots and walking parameters were analyzed automatically by FLLIT software. All the groups were repeated at least 3 times.

**Propidium Iodide and DAPI dual-stain in adult FeCO neurons**

The dual-staining protocol for PI and DAPI was adapted from the method described in (*53*, *54*). To perform dual-staining *in vivo*, we dissected the adult fly leg and incubated it without fixation in a solution containing 5 µg/ml Propidium Iodide (PI) (Invitrogen #00-6990) and a relatively high concentration of DAPI (1:20), allowing for the specific labeling of dead cells and the nuclei of FeCO neurons, respectively. Subsequently, the staining dye was diluted in PBS and incubated at 4°C for 1 day, followed by three 10-minute washes in PBS. Finally, the samples were mounted in VECTASHIELD with DAPI (Vector Laboratories #H-1200) for further analysis.

**RNA extraction and Real-time quantified PCR**

Total RNA was extracted from both human and fly samples using the NucleoSpin RNA Kit (Macherey-Nagel), and RNA concentration was normalized prior to cDNA synthesis. Reverse transcription was performed using the MMLV high-performance cDNA synthesis kit (Epicentre #ERT12910K). qRT-PCR was conducted using the IQ2 SYBR Green Fast qPCR System Master Mix (Bio Genesis #DBU-006) using an ABI QuantStudio 5 (Applied Biosystems). Human actin was used as the endogenous control for the human samples. All results represent the average of three independent experiments. The primers used were listed in table S6.

**Cell culture in SH-SY5Y cell**

**Generation of INSC stable knockdown cell line**

The pLKO.1 INSC shRNA plasmid, designed and provided by the RNA Technology Platform and Gene Manipulation Center of Academia Sinica in Taiwan, was used to generate a stable knockdown cell line of hINSC. The shRNA plasmid was purified using an Endotoxin-Free Midi-prep kit (Geneaid #PIFE25). Then, the shRNA and pGFP plasmids were co-transfected into SH-SY5Y cells for 24 hours. Prior to sorting, the cells were suspended in a sorting buffer with 1 μg/ml PI staining dye on ice. GFP-positive and PI-negative cells were sorted using a FACSAria IIIu flow cytometry sorter (BD bioscience), and single cells were sorted into 96-well plates in DMEM/F12 medium with 20% FBS. The sorted cells were incubated at 37°C for 2 weeks. A subset of the cells was harvested to validate the knockdown efficiency using real-time quantitative PCR, while the remaining cells were cultured in the 96-well plate for subsequent experiments.

**Propidium Iodide staining in SH-SY5Y cell**

For PI staining in SH-SY5Y cells, the cells were seeded onto cover glasses in 12 -well plate at a density of 1 x$10^5$ cells per well. After seeding for 24 hrs, the cells were stained with 1 ug/ml Propidium Iodide (PI)(Invitrogen #00-6990) diluted in PBS for 20 minutes at room temperature without fixation or light exposure. The stained cells were mounted in VECTASHIELD (Vector Laboratories #H-1200) with DAPI for further imaging.

**Transfection (plasmid)**

Human neuroblastoma SH-SY5Y cells were maintained at a density of 1 x$10^5$ cells per 12 -well plate in DMEM/F12 medium (Gibco #11320-033) containing 10% fetal bovine serum (FBS, Gibco #A3160502). Plasmids were transfected into cells by TransIT-X2® Dynamic Delivery System (Mirus Bio #MR-MIR6000) reagent for 24 hrs in the following transfection experiments.

**Immunostaining**

For acetyl-tubulin immunostaining, cells were seeded on cover glasses at a density of 1 x$10^5$ cells per 12 -well plate. The transfected cells were treated with DMSO, 0.8 ug/ml Taxol

(Sigma Aldrich #T1912) or 1 ug/ml Colchicine (Sigma Aldrich #C9754) for 6 hrs. After treatment, the cells were kept in DMEM/F12 for 24 hrs in 37$^{o}$C and then fixed with 4% paraformaldehyde (Electron Microscopy Sciences #15710) in PBS for 10 mins at room temperature. Fixed cells were incubated in 0.1% Triton-X (0.1% PBST) 10 minutes for cellular permeabilization and blocked with 1% BSA in PBS for 1 hrs in room temperature. After blocking, the cells were stained with anti-alpha-tubulin (1:500) (Abcam #ab6160), anti-acetyl-tubulin (1:500) (Sigma-Aldrich #T6793) and anti-FLAG tag (1:500) (Novus Biologicals #NBP1-06712) diluted in the blocking buffer overnight in 4$^{o}$C. On the next day, the primary antibodies were removed and cells washed for 5 mins 3 times with 0.4% Tween-20 (0.4% PBST) and incubated in secondary antibody for 1 hr at room temperature. The stained cells were mounted in the VECTASHIELD (Vector Laboratories #H-1200) with DAPI for further imaging. The catalog number and the working concentration of both primary and secondary antibodies were described in table S7.

**Neurite length quantification**

For the measurement of neurite length, the cells were seeded on cover glasses at a density of 1 x$10^5$ cells per 12-well plate. Subsequently, transfection and immunostaining of the cell were performed following standard procedures. The cell edge was determined with phalloidin (Invitrogen #A12380) staining.

**Quantification and statistical analysis**

The number of sensory neurons in the taris and the aggregative tubulin in the femur were quantified and analyzed by Imaris software in three-dimension images.
Quantitative data were analyzed using a two-tailed unpaired Student's *t-test* and the graphs were generated using GraphPad Prism 8. All data in graphs were expressed as mean±SEM. P values of less than 0.05 were considered statistically significant. * $p < 0.05$, ** $p < 0.01$, and *** $p < 0.001$. All of the statistical details of experiments can be found in the figure legends. n≥3 for each experiment with ≥3 independent experiments.

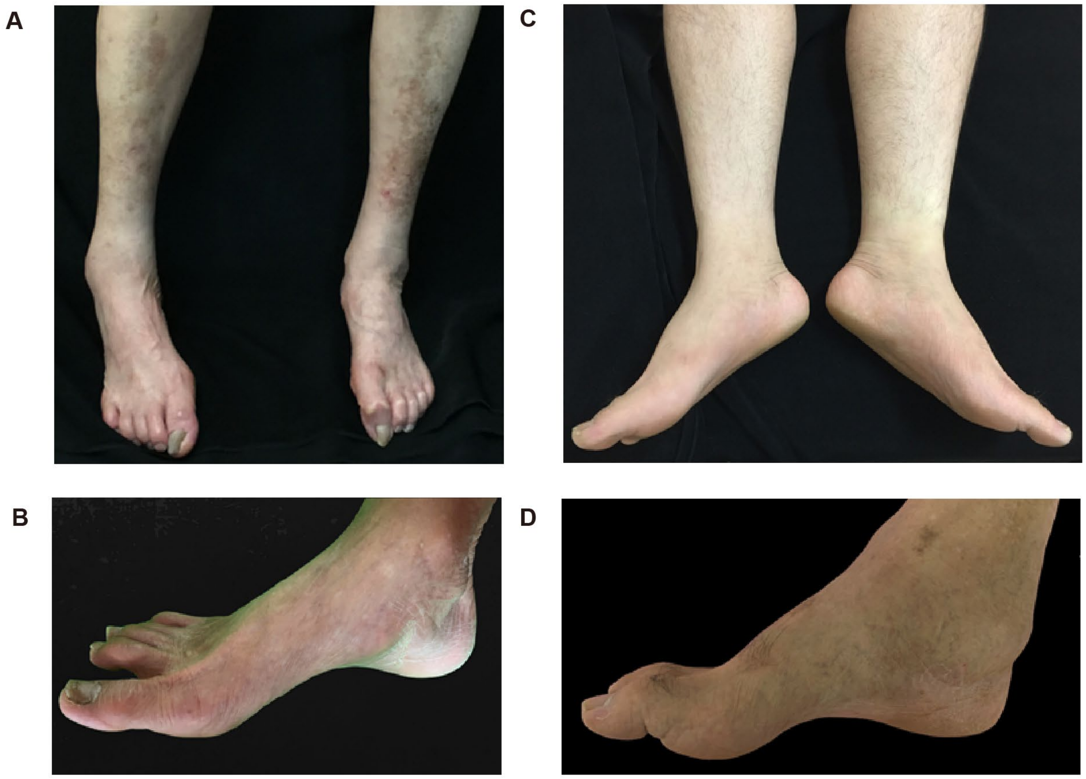


**Fig. S1. Clinical pictures of the patients with *INSC* p.Met70Arg.**
**(A)** Affected female II-2 in her 50's, with muscle wasting below the knees.
**(B)** Affected male II-3 in his 50's, with pes cavus.
**(C)** Affected male III-3 in his 30's, with pes planus.
**(D)** Affected male II-9 in his 40's, with pes cavus.

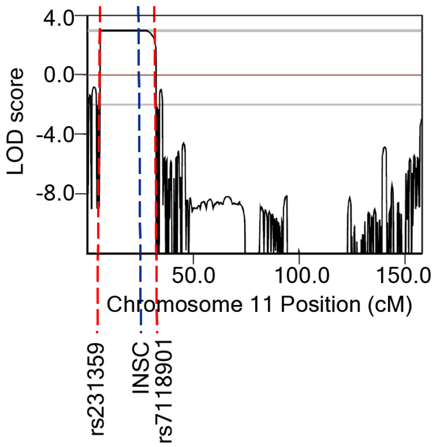


**Fig. S2**. **Multipoint parametric linkage analysis of CMT2 in the family.**
Multipoint parametric linkage analysis revealed three loci with a LOD score of approximately 3, including chr11:2694606-19743250 (GrCh37; Zmax 3.01), chr4:442021-6213622 (Zmax 3), and ch16:72022941-77978326 (Zmax 2.97). The red dotted lines define the flanking markers of the linkage peaks. The blue dotted line shows the location of the *INSC* gene within the chromosome 11 linkage region.

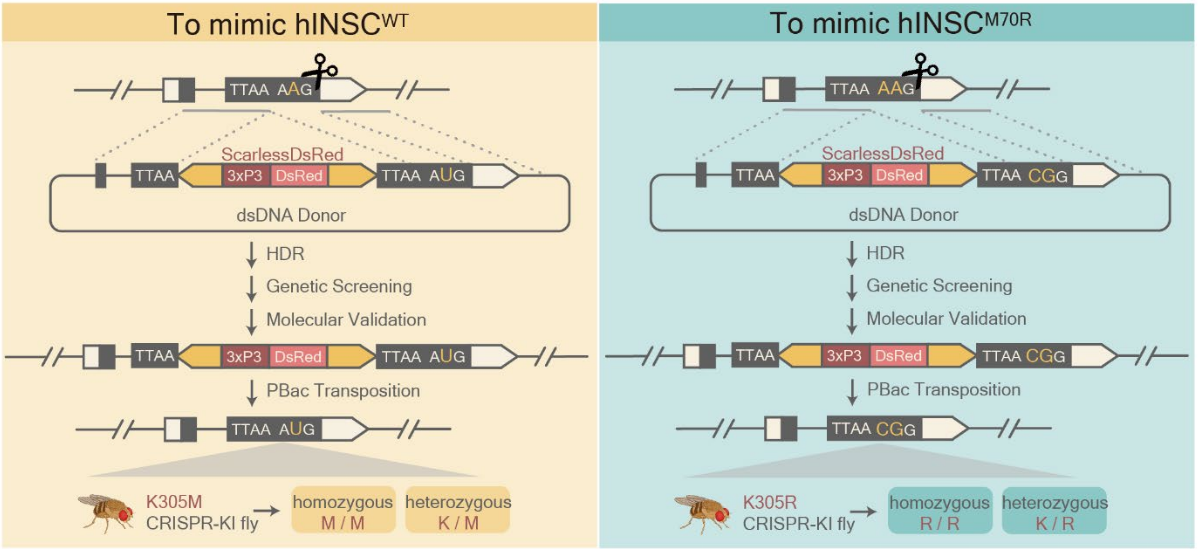


**Fig. S3**. **Schematic of a seamless editing knock-in strategy.**
Schematic presentation of a seamless editing knock-in strategy to generate two alleles, *dInsc*$^{K305M}$ and *dInsc*$^{K305R}$ with CRISPR-Cas9 technique to mimic *hINSC*$^{WT}$ and *hINSC*$^{M70R}$, respectively.

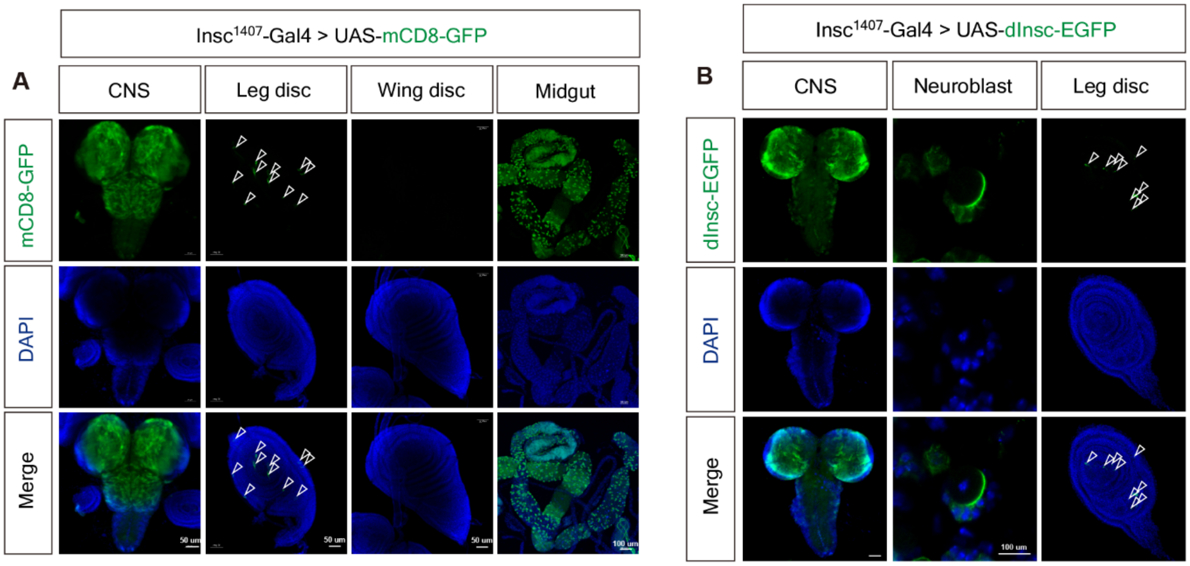


**Fig. S4. The expression pattern and protein localization of drosophila *Inscuteable* in larval tissues.**

**(A)** Representative images of CNS, leg disc, wing disc and midgut labeled with *mCD8*-GFP (green) under the control of *dInsc*$^{1407}$*-Gal4* in third instar larvae. Blue is DAPI staining. Scale bars: 50, 50, 50 and 100 µm from left to right.

**(B)** Representative confocal images of *dInsc-EGFP* (*dInsc*$^{1407}$*-Gal4>UAS-dInsc-EGFP*) in CNS, neuroblast, and leg disc of third instar larvae. Green indicates *dInsc*-EGFP. Blue is DAPI staining. Scale bars: 50, 100 and 50 µm from left to right.

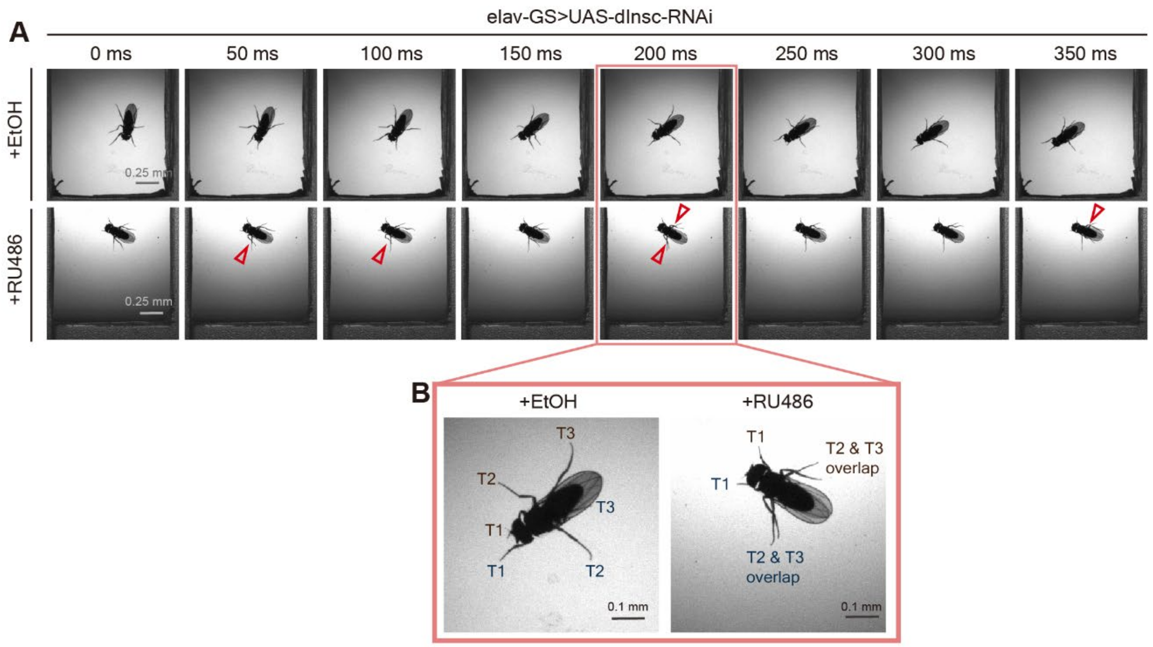


**Fig. S5. The time-lapse imaging of the *Drosophila* walking behaviors.**

**(A)** Snapshots from Drosophila in vivo imaging dataset for *elav-GS*>UAS-*dInsc*-RNAi with RU486 feeding (bottom row), compared with the solvent (EtOH)-feeding controls (top row). The interval of time-lapse is 50 ms. Scale bars: 0.25 mm

**(B)** Magnified images of (A). The RU486-fed flies, but not in solvent-feeding control, showed overlapped T2 and T3 legs (red arrowheads in (A)). Scale bars: 0.1 mm

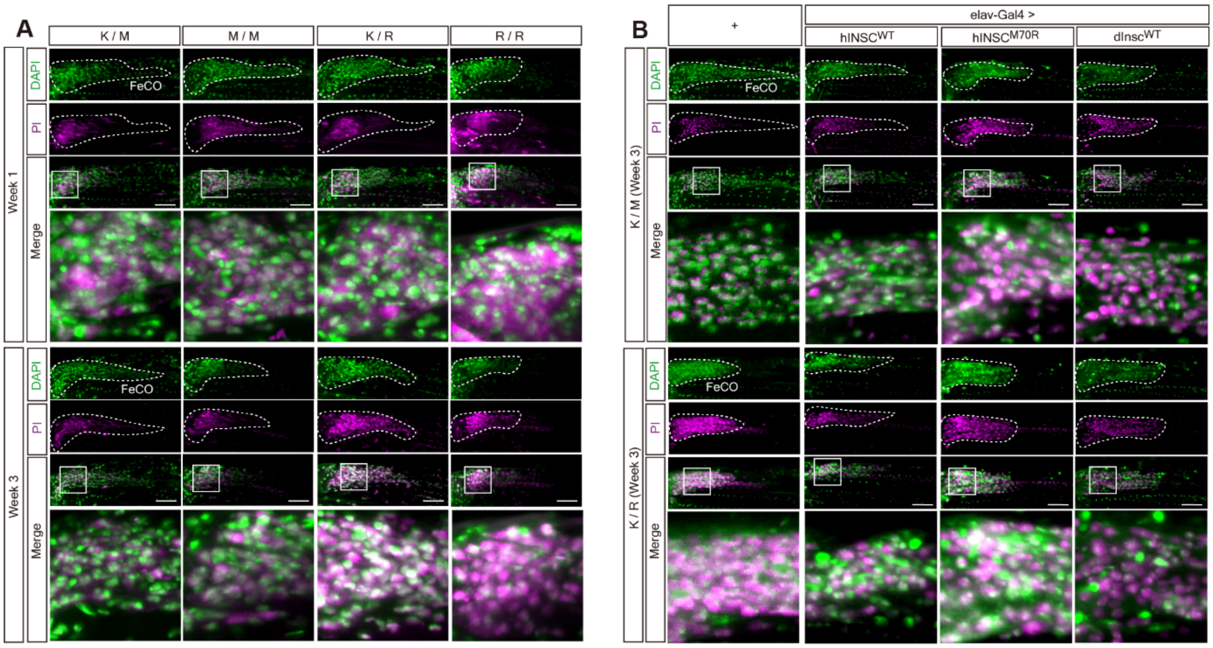


**Fig. S6. Representative confocal images of DAPI and PI co-staining in K305M and K305R CRISPR-KI flies.**

**(A)** Representative confocal images of $dInsc^{+/K305R}$ (*K/R*) and $dInsc^{K305R/K305R}$ (*R/R*) co-staining with PI (magenta) and DAPI (green) in FeCO neurons of 1- and 3-week-old flies, compared with $dInsc^{+/K305M}$ (*K/M*) and $dInsc^{K305M/K305M}$ (*M/M*) CRISPR-KI flies. The FeCO neurons are encircled by the dashed line. Scale bar: 5 µm

**(B)** Representative confocal images of overexpressing $hINSC^{WT}$, $hINSC^{M70R}$ and $dInsc^{WT}$ under the control of pan-neuronal *elav*-Gal4 in *K/M* and *K/R* background co-staining with PI (magenta) and DAPI (green) in FeCO neurons, comparing with 3-week-old *K/M* and *K/R* CRISPR-KI flies, respectively. The FeCO neurons are encircled by the dashed line. Scale bar: 5 µm

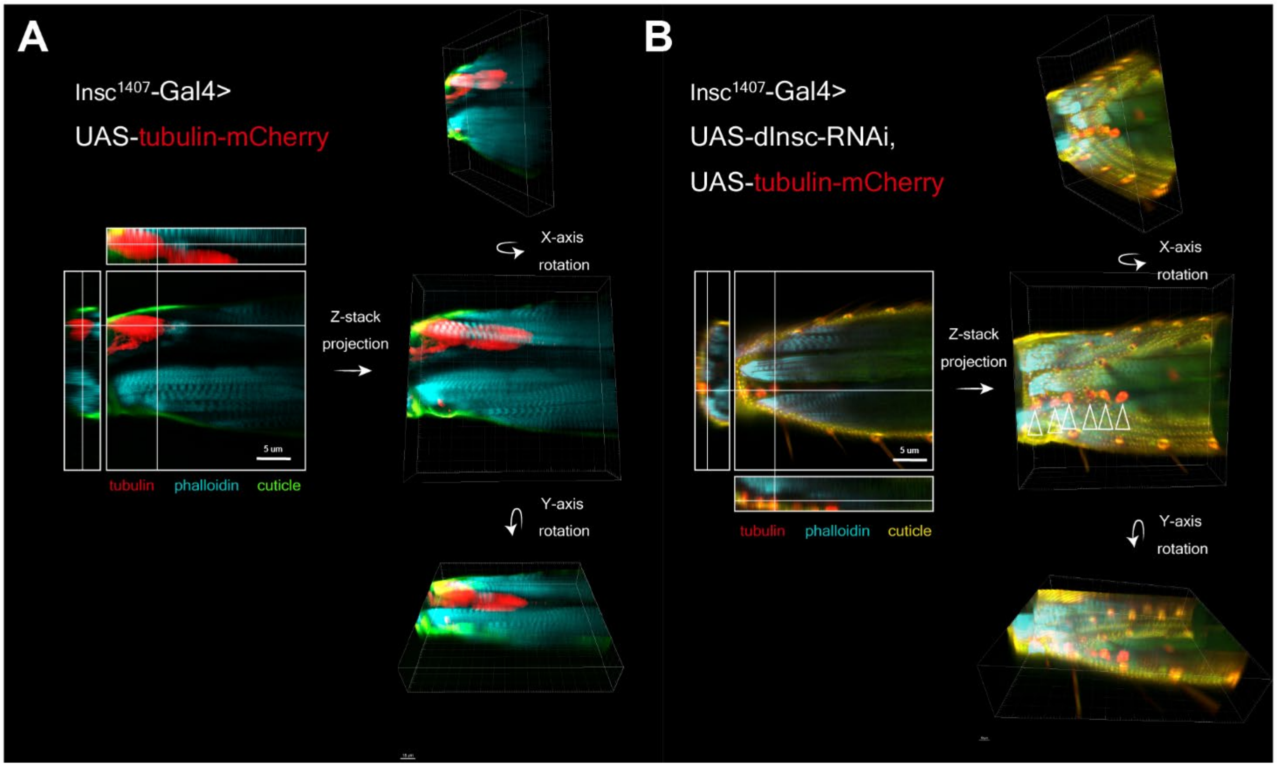


**Fig. S7. Three-dimensional imaging of tubulin aggregation in between muscle fibers in the femur.**
**(A, B)** Three-dimensional imaging of the femur of *dInsc*-RNAi flies under *dInsc$^{1407}$*-Gal4 compared with control (*tubulin-mCherry*) shows tubulin aggregation between muscle fibers. Red labeled tubulin, cyan labeled phalloidin (muscle fibers) and green indicated the auto-fluorescence of the cuticle. Arrowheads indicate the aggregative tubulin. Scale bars: 5 µm

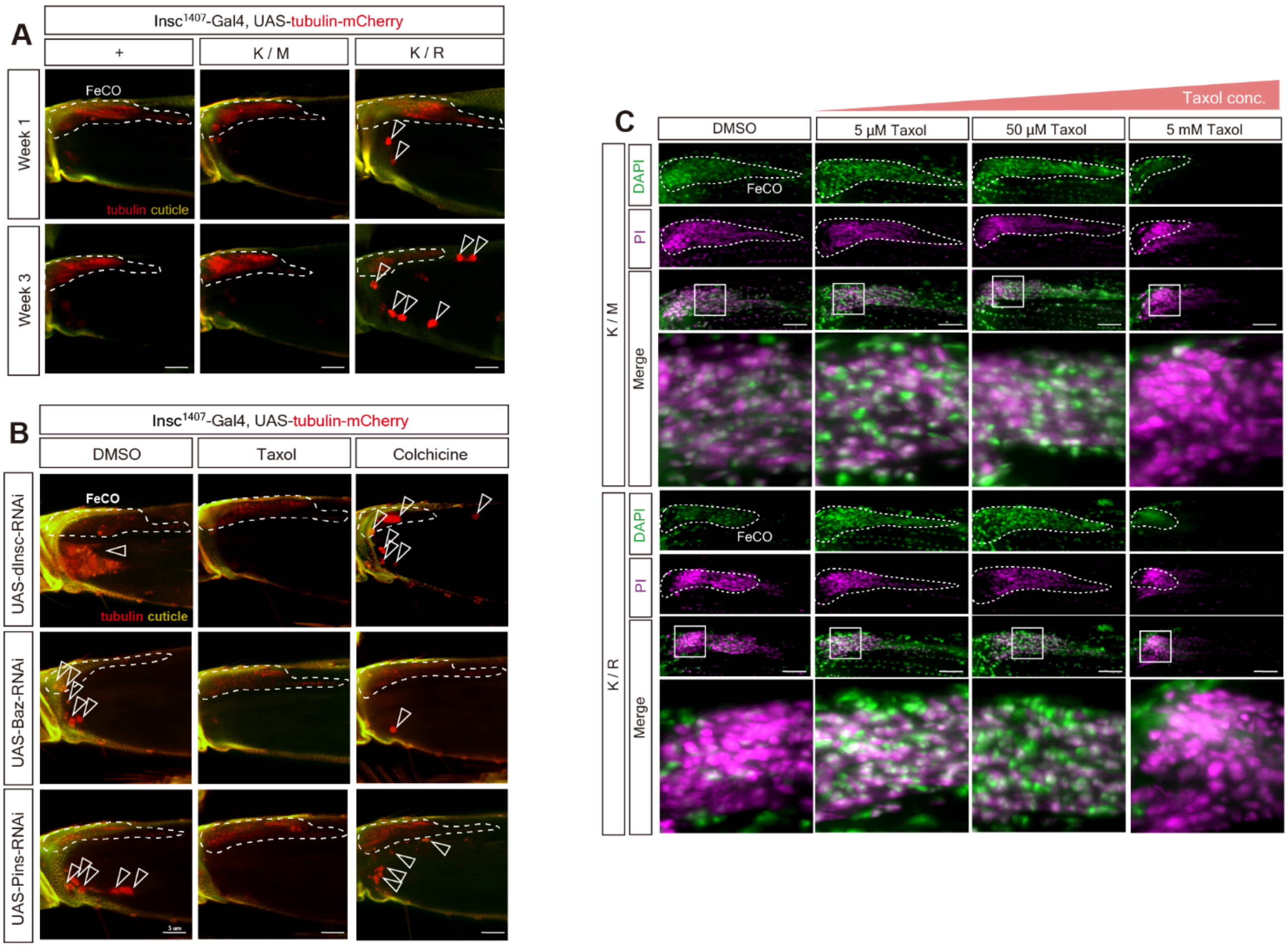


**Fig. S8. Taxol and colchicine exert opposite effects on tubulin accumulation.**

**(A)** Representative confocal images of tubulin aggregation in *K/M* and *K/R* of 1- and 3-week-old CRISPR-KI flies with *dInsc*$^{1407}$>UAS-*tubulin-mCherry*.

**(B)** Representative confocal images of tubulin aggregation (Red indicates *tubulin*-mCherry) in the femur of 3-week-old *dInsc*-RNAi, *Baz*-RNAi and *Pins*-RNAi flies under *dInsc*$^{1407}$-Gal4. Flies were treated with DMSO (vehicle control), Taxol (microtubule stabilizing agent) and Colchicine (microtubule destabilizing agent). Yellow indicated the auto-fluorescence of cuticles. The FeCO neurons are encircled by the dashed line. Arrowheads indicate the aggregative tubulin. Scale bars: 5 µm

**(C)** Representative confocal images of *K/M* and *K/R* CRISPR-KI flies treat with 3 different concentrations (5 µM, 50 µM, 5 mM) of microtubule stabilizer Taxol for 7 days co-staining with PI (magenta) and DAPI (green) in FeCO neurons, comparing with vehicle control (DMSO). The FeCO neurons are encircled by the dashed line. Scale bars: 5 µm

**Table. S1. Nerve conduction studies of the patients carrying *INSC* p.Met70Arg**

| Patient | Age at exam (y) | Motor nerve conduction studies | | | | | | Sensory nerve conduction studies | | | |
|---|---|---|---|---|---|---|---|---|---|---|---|
| | | Median nerve | | Peroneal nerve | | Tibial nerve | | Median nerve | | Sural nerve | |
| | | CV, m/s | CMAP, mV | CV, m/s | CMAP, mV | CV, m/s | CMAP, mV | CV, m/s | SNAP, uV | CV, m/s | SNAP, uV |
| II-2 | 55-60 | 53.9 | 6 | 37.6 | 0.3 | NR | NR | NR | NR | NR | NR |
| II-3 | 55-60 | 44.4 | 4.4 | ND | ND | ND | ND | NR | NR | ND | ND |
| II-8 | 51-55 | 56.6 | 4.7 | 45 | 2.6 | 42.2 | 2.0 | NR | NR | NR | NR |
| II-9 | 46-50 | 51.5 | 7 | 38 | 3.9 | 36.8 | 4.4 | 45.2 | 6 | NR | NR |
| III-3 | 26-30 | 55.5 | 12.2 | 39.6 | 2.1 | 40.1 | 1.4 | NR | NR | NR | NR |
| III-7 | 21-25 | 51.3 | 2.7 | 43.3 | 3.0 | 31.7 | 1.6 | NR | NR | NR | NR |
| III-8 | 21-25 | 54.5 | 5 | 41.7 | 0.7 | 36.8 | 0.7 | NR | NR | NR | NR |
| III-10 | 21-25 | 49 | 15.6 | 41.1 | 1.8 | 39.3 | 1 | NR | NR | NR | NR |
| Controls (mean ± SD) | | 58.6 ± 4 | 12.0 ± 3.9 | 48.8 ± 3.9 | 6.5 ± 2.9 | 49.6 ± 4.5 | 14.1 ± 4.5 | 56 ± 9 | 37.6 ± 16.4 | 48.3 ± 10.1 | 13.5 ± 6.6 |

Control values are based on a previously published report.[ref]

CV = conduction velocity; CMAP = compound motor action potential; SNAP = sensory action potential; NR = no response; ND = not done; SD = standard deviation.

Ref: Lin KP, Chan MH, Wu ZA. Nerve conduction studies in healthy Chinese: correlation with age, sex, height and skin temperature. Zhonghua Yi Xue Za Zhi (Taipei) 1993;52:293-297.

**Table. S2. Bioinformatics analyses and filtering of the whole genome sequencing data**

| Filtering criteria | Affected individuals with CMT | |
|---|---|---|
| | III-3 (S009) | III-8 (Q217) |
| Total bases sequenced by whole genome sequencing (Mbases) | 118,721 | 120,758 |
| Average depth in targeted region (x) | 37.2 | 38 |
| Percentage of >10x coverage in targeted region (%) | 99.5 | 99.6 |
| Heterozygous variants | 30,671 | 34,709 |
| Shared heterozygous variants in affected individuals | 14824 | |
| Variants not present in 1517 Taiwan biobank control genomes | 82 | |
| Variants altering coding sequences | 1 | |
| Variants completely segregated with CMT phenotype | 1 (*INSC* c.209T>G p.Met70Arg) | |

CMT: Charcot-Marie-Tooth disease

| Name | Genotype | Source |
|---|---|---|
| Insc$^{1407}$-Gal4 | *w[*]; P{w[+mW.hs]=GawB}insc[Mz1407]* | BDSC_8751 |
| UAS-dInsc-EGFP | *w[*]; P{UAS-dInsc.EGFP}2* | This study |
| UAS-mCD8-GFP | *y[1] w[*]; Pin[Yt]/CyO; P{w[+mC]=UAS-mCD8::GFP.L}LL6* | BDSC_5130 |
| elav-GS-Gal4 | *y[1] w[*]; P{w[+mC]=elav-Switch.O}GSG301* | BDSC_43642 |
| UAS-dInsc-RNAi | *y[1] v[1]; P{y[+t7.7] v[+t1.8]=TRiP.HMJ22537}attP40/CyO* | BDSC_60358 |
| UAS-w-RNAi | *y[1] v[1]; P{y[+t7.7] v[+t1.8]=TRiP.HMS00017}attP2* | BDSC_33623 |
| UAS-LacZ | *y[1] w[1118]; P{w[+mC]=UAS-lacZ.Exel}3* | BDSC_8530 |
| UAS-Pins-RNAi | *y[1] sc[*] v[1] sev[21]; P{y[+t7.7] v[+t1.8]=TKO.GS00670}attP40* | BDSC_77190 |
| UAS-Bazooka-RNAi | *y[1] v[1]; P{y[+t7.7] v[+t1.8]=TRiP.JF01079}attP2* | BDSC_31523 |
| W1118 | *W[1118]* | BDSC_3605 |
| Insc$^{InSITE}$-Gal4 | *w[1118]; PBac{w[+mC]=IT.GAL4}insc[1235-G4] sktl[1235-G4]/CyO* | BDSC_65626 |
| UAS-dInsc-WT | *w[*]; P{w[+mC]=UAS-insc.K}3* | BDSC_39676 |
| Iav-Gal4 | *w[*]; P{w[+mC]=iav-GAL4.K}3* | BDSC_52273 |
| OK371-Gal4 | *w[1118]; P{w[+mW.hs]=GawB}VGlut[OK371]* | BDSC_26160 |
| Insc$^{p49}$ | *insc[P49]/CyO* | DGRC_108467 |
| UAS-hINSC-WT-FLAG | *w[*]; P{UAS-hINSC-WT.FLAG}3* | This study |
| UAS-hINSC-M70R-FLAG | *w[*]; P{UAS-hINSC-M70R.FLAG}3* | This study |
| nan-Gal4 | *w[*]; P{w[+mC]=nan-GAL4.K}2* | BDSC_24903 |
| UAS-dPins-mCherry | *w[*]; P{UAS-dPins-mCherry}3* | This study |
| UAS-hNSC-WT-EGFP | *w[*]; P{UAS-hINSC-WT.EGFP}3* | This study |
| UAS-hNSC-M70R-EGFP | *w[*]; P{UAS-hINSC-M70R.EGFP}3* | This study |
| UAS-Bazooka-mCherry | *w[*]; P{y[+t7.7] w[+mC]=UAS-baz::mCherry}attP40/CyO* | BDSC_65844 |
| Insc$^{1407}$-Gal4, UAS-tub-mCherry | *w[*]; P{w[+mW.hs]=GawB}insc[Mz1407]/CyO; P{w[+mC]=UAS-ChRFP-Tub}3* | BDSC_25773 |
| Elav-Gal4 | *P{w[+mC]=GAL4-elav.L}2/CyO* | BDSC_8765 |

| | | |
|---|---|---|
| PiggyBac | *w[*]; PBac{GFP[ECFP.3xP3]=5pBlueEye.hsp70-PBac\T}Dmel2* | BDSC_32174 |
| CRISPR dInsc K305M KI | *dInsc-K305M-KI* | This study |
| CRISPR dInsc K305R KI | *dInsc-K305R-KI* | This study |

**Table. S3. Drosophila stocks**

| **Figures** | **Abbreviated genotype** | **Full genotype** |
|---|---|---|
| Fig.2A | elav-GS-Gal4>UAS-mCD8-GFP | w[*];+/+;P{w[+mC]=elav-Switch.O}GSG301/P{w[+mC]=UAS-mCD8::GFP.L}LL6 |
| Fig.2A | elav-GS-Gal4>UAS-dInsc-RNAi | w[*];P{y[+t7.7] v[+t1.8]=TRiP.HMJ22537}2/+; P{w[+mC]=elav-Switch.O}GSG301 |
| Fig.2A | elav-GS-Gal4>UAS-w-RNAi | w[*];+/+;P{w[+mC]=elav-Switch.O}GSG301/P{y[+t7.7] v[+t1.8]=TRiP.HMS00017}3 |
| Fig.2A | elav-GS-Gal4>UAS-Pins-RNAi | w[*];P{y[+t7.7] v[+t1.8]=TKO.GS00670}2/+; P{w[+mC]=elav-Switch.O}GSG301/+ |
| Fig.2A | elav-GS-Gal4>UAS-Baz-RNAi | w[*];+/+;;P{w[+mC]=elav-Switch.O}GSG301/P{y[+t7.7] v[+t1.8]=TRiP.JF01079}3 |
| Fig.2B | dInsc-InSITE-Gal4> UAS-dInsc-WT | w[*]; PBac{w[+mC]=IT.GAL4}insc[1235-G4] sktl[1235-G4]/+;P{w[+mC]=UAS-insc.K}3/+ |
| Fig.2B | dInsc-InSITE-Gal4 / + | w[*]; PBac{w[+mC]=IT.GAL4}insc[1235-G4] sktl[1235-G4]/+ |
| Fig.2B | dInsc-InSITE-Gal4> UAS-hINSC-WT | w[*]; PBac{w[+mC]=IT.GAL4}insc[1235-G4] sktl[1235-G4]/+;P{UAS-hINSC-WT.FLAG}3/+ |
| Fig.2B | dInsc-InSITE-Gal4> UAS-hINSC-M70R | w[*]; PBac{w[+mC]=IT.GAL4}insc[1235-G4] sktl[1235-G4]/+;P{UAS-hINSC-M70R.FLAG}3/+ |
| Fig.2B | dInsc-InSITE-Gal4> UAS-dInsc-RNAi | w[*]; PBac{w[+mC]=IT.GAL4}insc[1235-G4] sktl[1235-G4]/P{y[+t7.7] v[+t1.8]=TRiP.HMJ22537}attP40 |
| Fig.2B | dInsc-InSITE-Gal4> UAS-dInsc-RNAi, UAS-hINSC-WT | w[*]; PBac{w[+mC]=IT.GAL4}insc[1235-G4] sktl[1235-G4]/P{y[+t7.7] v[+t1.8]=TRiP.HMJ22537}attP40;P{UAS-hINSC-WT.FLAG}3/+ |
| Fig.2B | dInsc-InSITE-Gal4> UAS-dInsc-RNAi, UAS-hINSC-M70R | w[*]; PBac{w[+mC]=IT.GAL4}insc[1235-G4] sktl[1235-G4]/P{y[+t7.7] |

| | | |
|---|---|---|
| | | *v[+t1.8]=TRiP.HMJ22537}attP40;P{UAS-hINSC-M70R.FLAG}3/+* |
| Fig.2D | *dInsc k305M/+* | w[*];dInsc k305M-KI/+ |
| Fig.2D | *dInsc k305M/dInsc k305M* | w[*];dInsc k305M-KI/dInsc k305M-KI |
| Fig.2D | *dInsc k305R/+* | w[*];dInsc k305R-KI/+ |
| Fig.2D | *dInsc k305R/dInsc k305R* | w[*];dInsc k305R-KI/dInsc k305R-KI |
| Fig.2E, fig,S4A&B | *insc1407-Gal4>UAS-mCD8-GFP* | w[*]; P{w[+mW.hs]=GawB}insc[Mz1407]/+ ; P{w[+mC]=UAS-mCD8::GFP.L}LL6/+ |
| Fig.2I | *Iav-Gal4 > UAS-mCD8-GFP* | w[*];+/+; P{w[+mC]=iav-GAL4.K}3/P{w[+mC]=UAS-mCD8::GFP.L}LL6 |
| Fig.2I | *Iav-Gal4 > UAS-dInsc-RNA* | w[*];P{y[+t7.7] v[+t1.8]=TRiP.HMJ22537}2/+; P{w[+mC]=iav-GAL4.K}3/+ |
| Fig.2I | *Iav-Gal4 > UAS-Baz-RNAi* | w[*];+/+; P{w[+mC]=iav-GAL4.K}3/P{y[+t7.7] v[+t1.8]=TRiP.JF01079}3 |
| Fig.2I | *Iav-Gal4 > UAS-Pins-RNAi* | w[*];+/P{y[+t7.7] v[+t1.8]=TKO.GS00670}2;P{w[+mC]=iav-GAL4.K}3/+ |
| Fig.2J-N | *elav-GS-Gal4>UAS-dInsc-RNAi* | w[*];P{y[+t7.7] v[+t1.8]=TRiP.HMJ22537}2/+; P{w[+mC]=elav-Switch.O}GSG301 |
| Fig.3D&E | *nan-Gal4>UAS-w-RNAi* | w[*]; P{w[+mC]=nan-GAL4.K}2/+;P{y[+t7.7] v[+t1.8]=TRiP.HMS00017}3/+ |
| Fig.3D&E | *nan-Gal4>UAS-dInsc-RNAi* | w[*]; P{w[+mC]=nan-GAL4.K}2/P{y[+t7.7] v[+t1.8]=TRiP.HMJ22537}2;+/+ |
| Fig.3D&E | *nan-Gal4>UAS-Pins-RNAi* | w[*]; P{w[+mC]=nan-GAL4.K}2/P{y[+t7.7] v[+t1.8]=TKO.GS00670}2;+/+ |
| Fig.3D&E | *nan-Gal4>UAS-Baz-RNAi* | w[*]; P{w[+mC]=nan-GAL4.K}2/+;P{y[+t7.7] v[+t1.8]=TRiP.JF01079}3/+ |
| Fig.3F, fig.S6A | *dInsc k305M/+* | w[*];dInsc k305M-KI/+ |

| | | |
|---|---|---|
| Fig.3F, fig.S6A | *dInsc k305M/dInsc k305M* | w[*];dInsc k305M-KI/dInsc k305M-KI |
| Fig.3F, fig.S6A | *dInsc k305R/+* | w[*];dInsc k305R-KI/+ |
| Fig.3F, fig.S6A | *dInsc k305R/dInsc k305R* | w[*];dInsc k305R-KI/dInsc k305R-KI |
| Fig.3G, fig.S6B | *dInsc k305M/+* | w[*];dInsc k305M-KI/+ |
| Fig.3G, fig.S6B | *dInsc k305M; elav-Gal4>UAS-hINSC-WT* | w[*];dInsc k305M-KI/P{w[+mC]=GAL4-elav.L}2;P{UAS-hINSC-WT.FLAG}3/+ |
| Fig.3G, fig.S6B | *dInsc k305M; elav-Gal4>UAS-hINSC-M70R* | w[*];dInsc k305M-KI/P{w[+mC]=GAL4-elav.L}2;P{UAS-hINSC-M70R.FLAG}3/+ |
| Fig.3G, fig.S6B | *dInsc k305M; elav-Gal4>UAS-dInsc-WT* | w[*];dInsc k305M-KI/P{w[+mC]=GAL4-elav.L}2;P{w[+mC]=UAS-insc.K}3/+ |
| Fig.3G, fig.S6B | *dInsc k305R/+* | w[*];dInsc k305R-KI/+ |
| Fig.3G, fig.S6B | *dInsc k305R; elav-Gal4>UAS-hINSC-WT* | w[*];dInsc k305R-KI/P{w[+mC]=GAL4-elav.L}2;P{UAS-hINSC-WT.FLAG}3/+ |
| Fig.3G, fig.S6B | *dInsc k305R; elav-Gal4>UAS-hINSC-M70R* | w[*];dInsc k305R-KI/P{w[+mC]=GAL4-elav.L}2;P{UAS-hINSC-M70R.FLAG}3/+ |
| Fig.3G, fig.S6B | *dInsc k305R; elav-Gal4>UAS-dInsc-WT* | w[*];dInsc k305R-KI/P{w[+mC]=GAL4-elav.L}2;P{w[+mC]=UAS-insc.K}3/+ |
| Fig.4B&C,I &J | *nan-Gal4>UAS-hINSC-WT, UAS-Pins-mCherry* | w[*]; P{w[+mC]=nan-GAL4.K}2/+;P{UAS-hINSC-WT.FLAG}3/P{UAS-dPins-mCherry}3 |
| Fig.4B&C,I &J | *nan-Gal4>UAS-hINSC-M70R, UAS-Pins-mCherry* | w[*]; P{w[+mC]=nan-GAL4.K}2/+;P{UAS-hINSC-M70R.FLAG}3/P{UAS-dPins-mCherry}3 |
| Fig.4H | *nan-Gal4>UAS-hINSC-WT, UAS-Baz-mCherry* | w[*]; P{w[+mC]=nan-GAL4.K}2/P{y[+t7.7] w[+mC]=UAS-baz::mCherry}2;P{UAS-hINSC-WT.FLAG}3/+ |

| | | |
|---|---|---|
| Fig.4H | *nan-Gal4>UAS-hINSC-M70R, UAS-Baz-mCherry* | w[*]; P{w[+mC]=nan-GAL4.K}2/P{y[+t7.7] w[+mC]=UAS-baz::mCherry}2;P{UAS-hINSC-M70R.FLAG}3/+ |
| Fig.5A-C | *Insc1407-Gal4> UAS-tubulin-mCherry, UAS-w-RNAi* | w[*]; P{w[+mW.hs]=GawB}insc[Mz1407]/+; P{w[+mC]=UAS-ChRFP-Tub}3/P{y[+t7.7] v[+t1.8]=TRiP.HMS00017}3 |
| Fig.5A-C | *Insc1407-Gal4> UAS-tubulin-mCherry, UAS-dInsc-RNAi* | w[*]; P{w[+mW.hs]=GawB}insc[Mz1407]/P{y[+t7.7] v[+t1.8]=TRiP.HMJ22537}2; P{w[+mC]=UAS-ChRFP-Tub}3/+ |
| Fig.5A-C | *Insc1407-Gal4> UAS-tubulin-mCherry, UAS-dInsc-RNAi, UAS-hINSC-WT* | w[*]; P{w[+mW.hs]=GawB}insc[Mz1407]/P{y[+t7.7] v[+t1.8]=TRiP.HMJ22537}2; P{w[+mC]=UAS-ChRFP-Tub}3/P{UAS-hINSC-WT.FLAG}3 |
| Fig.5A-C | *Insc1407-Gal4> UAS-tubulin-mCherry, UAS-dInsc-RNAi, UAS-hINSC-M70R* | w[*]; P{w[+mW.hs]=GawB}insc[Mz1407]/P{y[+t7.7] v[+t1.8]=TRiP.HMJ22537}2; P{w[+mC]=UAS-ChRFP-Tub}3/P{UAS-hINSC-M70R.FLAG}3 |
| Fig.5D&E, fig.S8A | *W1118* | w[1118] |
| Fig.5D&E, fig.S8A | *dInsc k305M/+* | w[*];dInsc k305M-KI/+ |
| Fig.5D&E, fig.S8A | *dInsc k305R/+* | w[*];dInsc k305R-KI/+ |
| Fig.5F&G, fig.S8B | *Insc1407-Gal4> UAS-tubulin-mCherry, UAS-dInsc-RNAi* | w[*]; P{w[+mW.hs]=GawB}insc[Mz1407]/P{y[+t7.7] v[+t1.8]=TRiP.HMJ22537}2; P{w[+mC]=UAS-ChRFP-Tub}3/+ |
| Fig.5F&G, fig.S8B | *Insc1407-Gal4> UAS-tubulin-mCherry, UAS-Baz-RNAi* | w[*]; P{w[+mW.hs]=GawB}insc[Mz1407]/+;P{y[+t7.7] v[+t1.8]=TRiP.JF01079}3/P{w[+mC]=UAS-ChRFP-Tub}3 |

| | | |
|---|---|---|
| Fig.5F&G, fig.S8B | *Insc1407-Gal4> UAS-tubulin-mCherry, UAS-Pins-RNAi* | w[*]; P{w[+mW.hs]=GawB}insc[Mz1407]/P{y[+t7.7] v[+t1.8]=TKO.GS00670}2;P{w[+mC]=UAS-ChRFP-Tub}3/+ |
| Fig.6A&D | *dInsc k305M; Insc1407-Gal4> UAS-tubulin-mCherry* | w[*];dInsc k305M-KI/P{w[+mW.hs]=GawB}insc[Mz1407];P{w[+mC]=UAS-ChRFP-Tub}3/+ |
| Fig.6A&D | *dInsc k305R; Insc1407-Gal4> UAS-tubulin-mCherry* | w[*];dInsc k305R-KI/P{w[+mW.hs]=GawB}insc[Mz1407];P{w[+mC]=UAS-ChRFP-Tub}3/+ |
| Fig.6H, fig.S8C | *dInsc k305M/+* | w[*];dInsc k305M-KI/+ |
| Fig.6H, fig.S8C | *dInsc k305R/+* | w[*];dInsc k305R-KI/+ |
| Fig.S7A | *Insc1407-Gal4> UAS-tubulin-mCherry* | w[*]; P{w[+mW.hs]=GawB}insc[Mz1407]/+; P{w[+mC]=UAS-ChRFP-Tub}3/+ |
| Fig.S7B | *Insc1407-Gal4> UAS-tubulin-mCherry, UAS-dInsc-RNAi* | w[*]; P{w[+mW.hs]=GawB}insc[Mz1407]/P{y[+t7.7] v[+t1.8]=TRiP.HMJ22537}2; P{w[+mC]=UAS-ChRFP-Tub}3/+ |

**Table. S4. *Drosophila* genotype and associated figures**

| **Name** | **Source** |
|---|---|
| pUASt-attB-hINSC$^{WT}$-FLAG | This study (FLAG tag fused to C terminus) |
| pUASt-attB-UAS-hINSC$^{M70R}$-FLAG | This study (FLAG tag fused to C terminus) |
| pUASt-attB-UAS-hINSC$^{WT}$-EGFP | This study (EGFP fused to C terminus) |
| pUASt-attB-UAS-hINSC$^{M70R}$-EGFP | This study (EGFP fused to C terminus) |
| pUASt-attB-UAS-dInsc-EGFP | This study (EGFP fused to C terminus) |
| pUASt-attB-UAS-Pins-mCherry | This study (mCherry fused to C terminus) |
| pFLAG-hINSC$^{WT}$ | This study (FLAG tag fused to C terminus) |
| pFLAG-hINSC$^{M70R}$ | This study (FLAG tag fused to C terminus) |
| pMyc-LGN | This study (Myc tag fused to C terminus) |
| pHA-PAR3 | This study (HA tag fused to C terminus) |
| pBFv-U6.2 | Addgene #138400 |
| pScarlessHD-DsRed | Addgene #64703 |

**Table. S5. List of plasmids used in this study**

| Quantitative real-time PCR primers | |
|---|---|
| **Gene** | **Primer sequence** |
| *human INSC* | fwd: ATGATGGCACTGCCTGGAG |
| | rev: CTGCAGGACACACATGCACT |
| *human Actin* | fwd: GCGTCGGTCAATTCAATCTT |
| | rev: AAGCTGCAACCTCTTCGTCA |
| **Site-Directed Mutagenesis primers** | |
| **Gene** | **Primer sequence** |
| *hINSC-M70R* | fwd: GCGGCTACACCTGAGGCAGGTGGACTCAGTC |
| | rev: GACTGAGTCCACCTGCCTCAGGTGTAGCCGC |
| **Molecular cloning primers** | |
| **Plasmid** | **Primer sequence** |
| pUASt-attB-UAS-dInsc-EGFP | fwd (BglII site): ATTCGTTAACAGATCTATGTCCTTTCAGCGTAGC<br>rev (XhoI site): TAGAGGTACCCTCGAGTTACTTGTACAGCTCGTCC |
| pUASt-attB-UAS-dPins-mCherry | fwd (EcoRI site): AGGGAATTGGGAATTCATGTCCTCGCTCTCTGCG |
| | rev (KpnI site): AAAGATCCTCTAGAGGTACCTTACTTGTACAGCTCGTCCAT |
| pUASt-attB-UAS-hINSC-WT-FLAG | fwd (EcoRI site): CCCGAATTCCAAAATGAGACGGCCCCCTGGC |
| | rev (XhoI site): GGGCTCGAGCTACTTGTCATCGTCGTC |
| pUASt-attB-UAS-hINSC-M70R-FLAG | fwd (EcoRI site): CCCGAATTCCAAAATGAGACGGCCCCCTGGC |
| | rev (XhoI site): GGGCTCGAGCTACTTGTCATCGTCGTC |
| pUASt-attB-UAS-hINSC-WT -EGFP | fwd (KpnI site): GTTTTGTGGGATCCGGTACCATGGTGAGCAAGGGCGAG |

| | |
|---|---|
| | rev (KpnI site): AAAGATCCTCTAGAGGTACCTTACTTGTACAGCTCGTCCA |
| pUASt-attB-UAS-hINSC-M70R - EGFP | fwd (KpnI site): GTTTTGTGGGATCCGGTACCATGGTGAGCAAGGGCGAG |
| | rev (KpnI site): AAAGATCCTCTAGAGGTACCTTACTTGTACAGCTCGTCCA |
| pFLAG-hINSC-WT | fwd (EcoRI site): AAATTGAATTCACCATGAGACGGCCCCC |
| | rev (BamHI site): CGCGGATCCCACAAAACTCTCCTCC |
| pFLAG-hINSC-M70R | fwd (EcoRI site): AAATTGAATTCACCATGAGACGGCCCCC |
| | rev (BamHI site): CGCGGATCCCACAAAACTCTCCTCC |

**Table. S6. List of primers used in this study**

| Name | Source | Identifier | concentration |
|---|---|---|---|
| Rat anti-FLAG | Novus biologicals | Novus Cat# NBP1-06712; RRID:AB_1625981 | 1:500 |
| Mouse anti-DLG | DSHB | DSHB Cat# 4F3 | 1:200 |
| Rabbit anti-HA | Abcam | Abcam Cat# ab9110; RRID:AB_307019 | 1:500 |
| Mouse anti-c-Myc | DSHB | DSHB Cat# 9E10; RRID:AB_2266850 | 1:500 |
| Rat anti-alpha-tubulin | Abcam | Abcam Cat# ab6160; RRID:AB_305328 | 1:500 |
| Mouse anti-acetylated tubulin | Sigma-Aldrich | Sigma-Aldrich Cat# T6793; RRID:AB_477585 | 1:500 |
| Alexa Fluor@-568 Phalloidin | Invitrogen | Thermo Fisher Scientific Cat#A12380 | 1:500 |
| Alexa Fluor@-488 anti-Mouse IgG | Jackson ImmunoResearch Laboratories | Jackson ImmunoResearch Labs Cat# 715-545-150; RRID:AB_2340846 | 1:500 |

**Table. S7. List of antibodies used in this study**

**Movie. S1.**
The video of the patient II-3 in his 50's demonstrating a wide-based steppage gait, suggesting foot drop with sensory ataxia.

**Movie. S2.**
*In vivo* imaging of *Insc* knockdown flies treated with EtOH.

**Movie. S3.**
*In vivo* imaging of *Insc* knockdown flies treated with RU486.
